# Alternative nutritional and clinical care practices for NEC prevention drive distinct profiles and functional responses in the preterm gut microbiome

**DOI:** 10.1101/2022.06.08.22276177

**Authors:** Charlotte J Neumann, Alexander Mahnert, Christina Kumpitsch, Raymond Kiu, Matthew J Dalby, Magdalena Kujawska, Tobias Madl, Stefan Kurath-Koller, Berndt Urlesberger, Bernhard Resch, Lindsay J Hall, Christine Moissl-Eichinger

## Abstract

Preterm infants with very low birthweight are at serious risk for necrotizing enterocolitis. To functionally analyse the principles of three successful preventive NEC regimens, we characterized faecal samples of 54 infants (< 1,500 g, n = 383) longitudinally (two weeks) with respect to gut microbiome profiles (bacteria, archaea, fungi, viruses), microbial function, virulence factors, antibiotic resistances and metabolic profiles, including human milk oligosaccharides (HMOs) and short-chain fatty acids. Probiotic *Bifidobacterium longum* ssp. *infantis* supplementation affected microbiome development globally, pointing toward the genomic potential to convert HMOs. Engraftment of *Bifidobacterium* substantially reduced microbiome-associated antibiotic resistance as compared to regimens using probiotic *Lactobacillus rhamnosus* or no supplementation. Crucially, the beneficial effects of *Bifidobacterium* supplementation depended on simultaneous feeding with HMOs. We demonstrate that preventive regimens have the highest impact on early maturation of the gastrointestinal microbiome, enabling the establishment of a resilient microbial ecosystem that reduces pathogenic threats in at-risk preterm infants.

## Introduction

About eleven percent of all infants worldwide are born prematurely, i.e. before 37 weeks’ gestation (Blencowe et al., 2012). Very low birth weight (VLBW) preterm infants (< 1,500 g) are particularly vulnerable to acute and long-term clinical complications. Of particular concern is the development of necrotizing enterocolitis (NEC), a serious gastrointestinal threat that occurs in 7–11% of VLBW infants (Fanaroff et al., 2007). In such cases, mortality can reach up to 30% (Neu and Walker, 2011).

NEC is a devastating multifactorial disease that is driven in part by perturbations of the microbiome, including colonization and overgrowth of certain microbes with potentially pathogenic potential such as *Escherichia coli* or *Clostridium perfringens* (Högberg et al., 2013).

Given the rapid onset of NEC, a number of neonatal intensive care units (NICUs) have developed specific NEC prophylaxis programmes that include the use of probiotics, antibiotics and differentiated feeding protocols and that have resulted in a recent, substantial decrease in NEC rates in preterm infants (Jin et al., 2019).

Probiotic treatments are usually based on the use of *Bifidobacterium* and *Lactobacillus* species (Alcon-Giner et al., 2020). *Bifidobacterium*, in particular, is considered as an important member of the resident infant microbiome that is maintained into early childhood and promotes healthy infant development (Bäckhed et al., 2015; Oki et al., 2018).

Antibiotics are administered intravenously at the first signs of infection to control early-onset sepsis. As a result, the majority of VLBW infants are exposed to antibiotics in the first few days of life and for extended periods of time (Bizzarro, 2018). A number of publications have emphasized the need for responsible antibiotic usage in such vulnerable patients, as their use is associated with the risk of infection with multi-drug resistant (MDR) pathogens such as *Staphylococcus aureus*, *Escherichia coli* and *Klebsiella pneumoniae* (van Duin and Paterson, 2016) and is believed to have other, largely unknown, long-term effects. Overall, antibiotic exposure is often considered as preventable (Bizzarro, 2018; Gustavsson et al., 2020).

Additionally, the use of enteral antibiotics may be effective as NEC prophylaxis. The Cochrane Neonatal Collaborative Review Group (Bury and Tudehope, 2001) evaluated five trials where oral antibiotics were used as a prophylaxis against NEC in low-birth-weight preterm infants. Their findings suggest that oral administration of prophylactic enteral antibiotics results in a statistically significant reduction in NEC and in NEC-related deaths. However, the risks of enteral antibiotics have not yet been quantified; thus, this strategy has never been widely adopted due to concerns about the emergence of resistant bacteria and the absorption of antibiotics from the gut (Bell, 2005). However, such adverse effects have not been reported so far (Schmolzer et al., 2006).

Human milk (HM), the gold standard for infant feeding, is a surprisingly complex synbiotic that contains probiotic bacteria and prebiotics to nourish probiotic bacteria. Prebiotic human milk oligosaccharides (HMOs) are complex carbohydrates present in large quantities in HM which are not broken down by intestinal enzymes. Therefore, they serve only as a specific substrate for certain bacteria in the infant’s gastrointestinal tract (GIT), such as mainly *Bifidobacterium* (*Bifidobacterium bifidum* and *Bifidobacterium longum* subsp. *infantis*) but also *Bacteroides* (*Bacteroides vulgatus* and *Bacteroides fragilis*) (Kujawska et al., 2021; Marcobal and Sonnenburg, 2012). Indeed, *Bifidobacterium* is enriched in infants fed HM (Ruiz et al., 2019), due to its ability to metabolize HMOs. The sophisticated, individual complexity of HM can only be partially mimicked by formula milk (FM); nevertheless, newer products also contain standardized pre- and probiotics for optimal nutrition.

Southern Austrian neonatal units have implemented various combinations of these prophylactic measures with great success, resulting in an exceptionally low average NEC rate of 2.9% in VLBW infants (2007-2016; (Wellmann, 2018)). We took the opportunity in this study to deeply analyse the mechanism for success across these different regimens on the level of the gut microbiome and metabolome.

We recruited 54 VLBW infants in three hospital centres (Graz, Klagenfurt, Leoben), that differed in antibiotic treatment (enteral gentamicin or none), antifungal treatment (enteral nystatin or parenteral fluconazole), probiotic use (*Bifidobacterium*, *Lactobacillus*, or none) and feeding (HM, FM). Using a multi-omics approach, we examined the composition and function of the microbiome and its metabolites in the first weeks of life to understand the importance of the interactions among dietary components, antibiotics and probiotics.

Our study differs from previous studies in that a focus was placed on successful but different NEC-prevention protocols, not in just one but in three different clinics. This study setup also allowed us to avoid the problematic cross-contamination of probiotics into the control groups (Costeloe et al., 2016; Karthikeyan and Bhat, 2017). To understand the effects and mechanisms of the different treatments, we analysed the microbiome on a multi-kingdom level and included functional metagenomics and metabolomics, as well as genome profiling on the species level. We conclude our study with a suggestion to further improve existing protocols to support a healthy microbiome development in VLBW infants by combining effective probiotics (including *Bifidobacterium*) and human milk.

## Results

Details on the study design are provided within the Star Methods. In brief, faecal samples were collected prospectively in three independent NICUs in Austria using a different NEC prophylaxis regimen (**Table 1**, in Methods) from preterm infants with a birthweight < 1,500 g. Samples were collected every other day, starting with the meconium, up until two weeks of age.

**Table 1:**
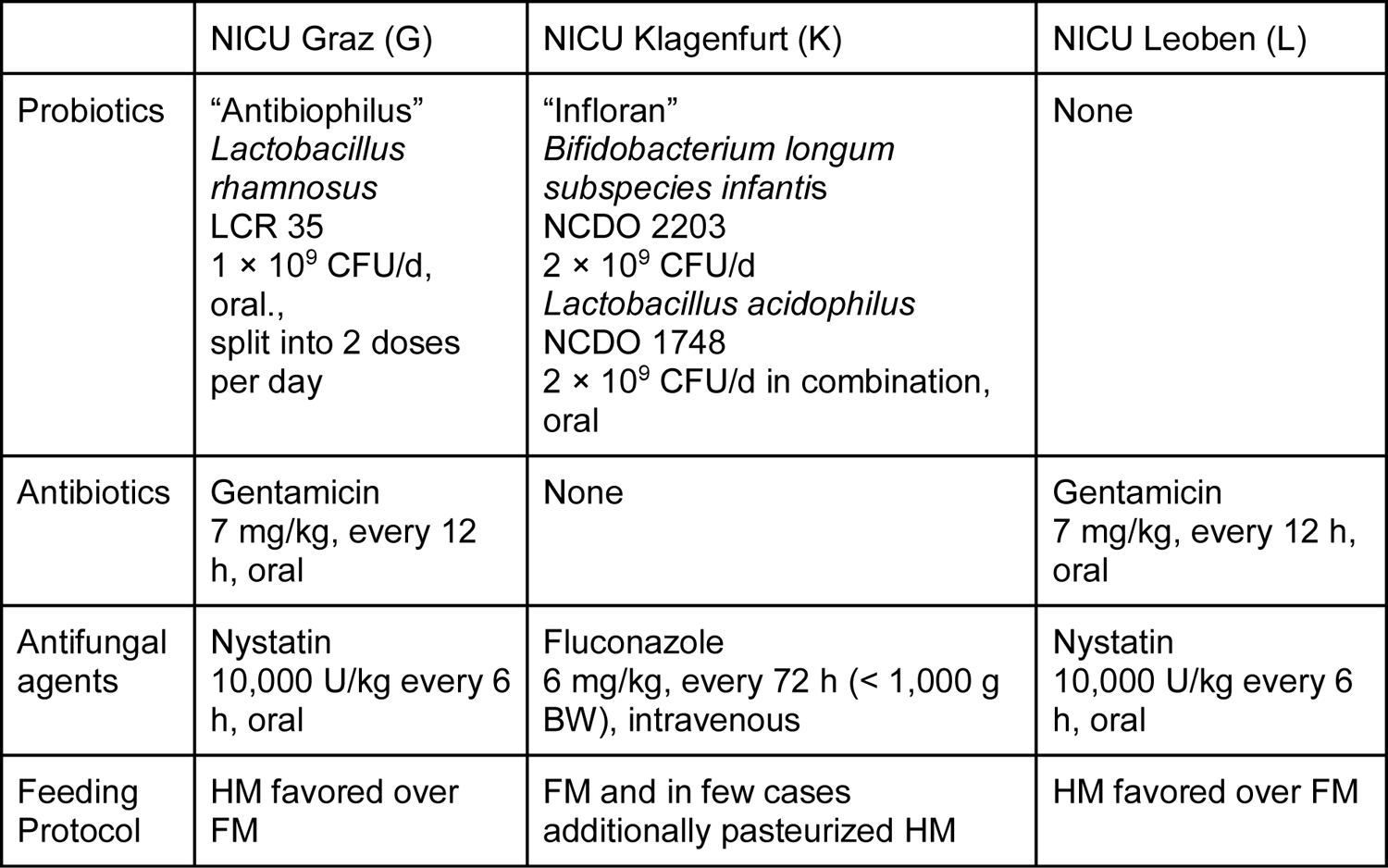
prophylactic regimens of probiotics, antibiotics, antifungals and feeding protocols of the three different neonatal intensive care units (NICUs) Graz (G), Klagenfurt (K) and Leoben (L). CFU: Colony forming units; HM: Human milk; FM: Formula milk

### Early-life therapy regimen influences microbiome composition and development across all microbial domains

Assessing the microbial composition of all infants with metagenomic analyses and 16S rRNA gene sequencing, we detected a global effect of the preventive NEC regimen on <u>all</u> microbial domains and groups, including bacteria (99.62% of all metagenomic reads), their phages and viruses (0.15%), but also on archaea (0.04%) and fungi (Ascomycota/ Basidiomycota: 0.09%) (**Table 2, Fig. 1, Fig. 2**).

**Table 2:**
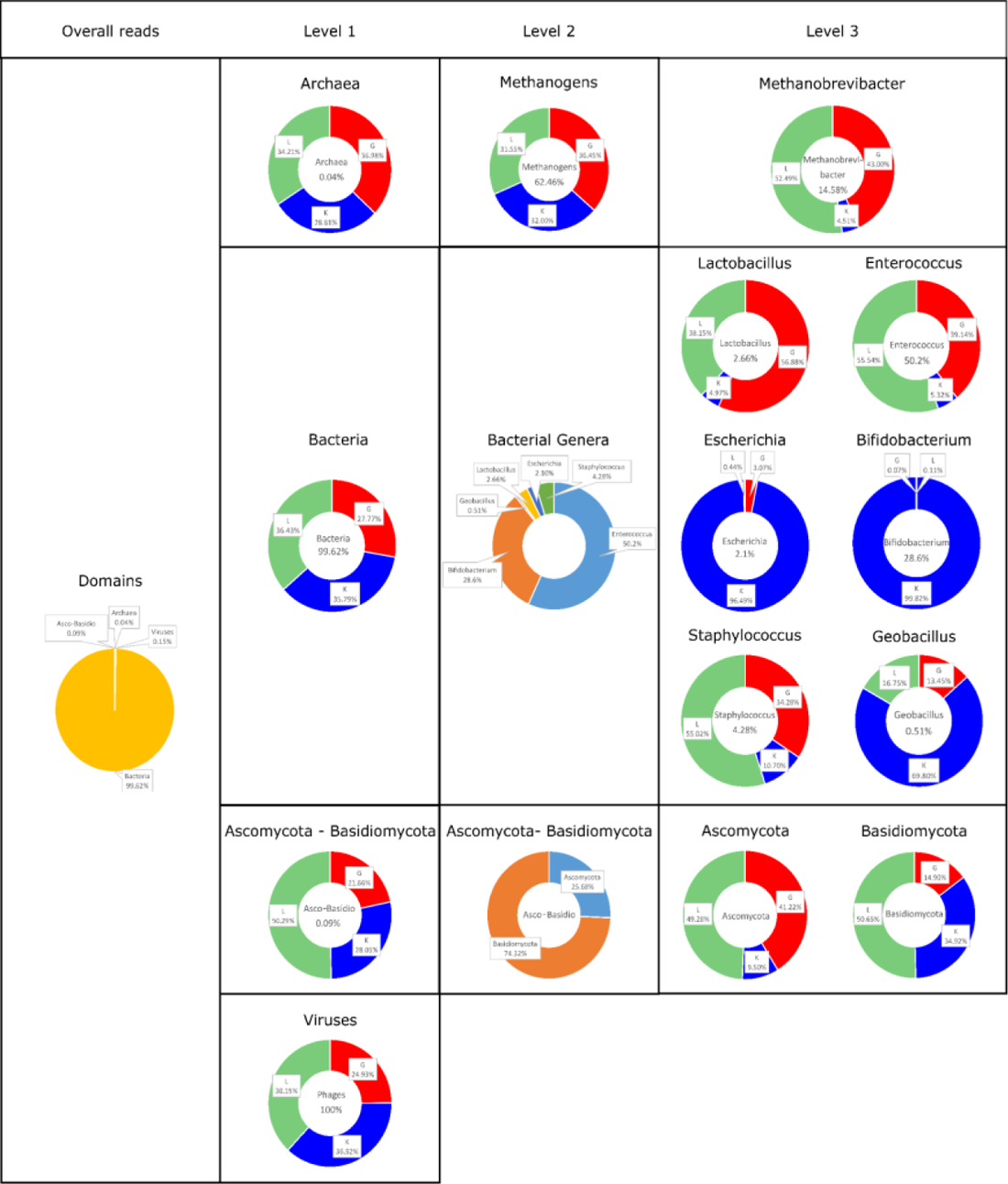
Distribution of overall metagenomic reads across the domains of life and between the centres on different taxonomic levels.

**Figure 1:**
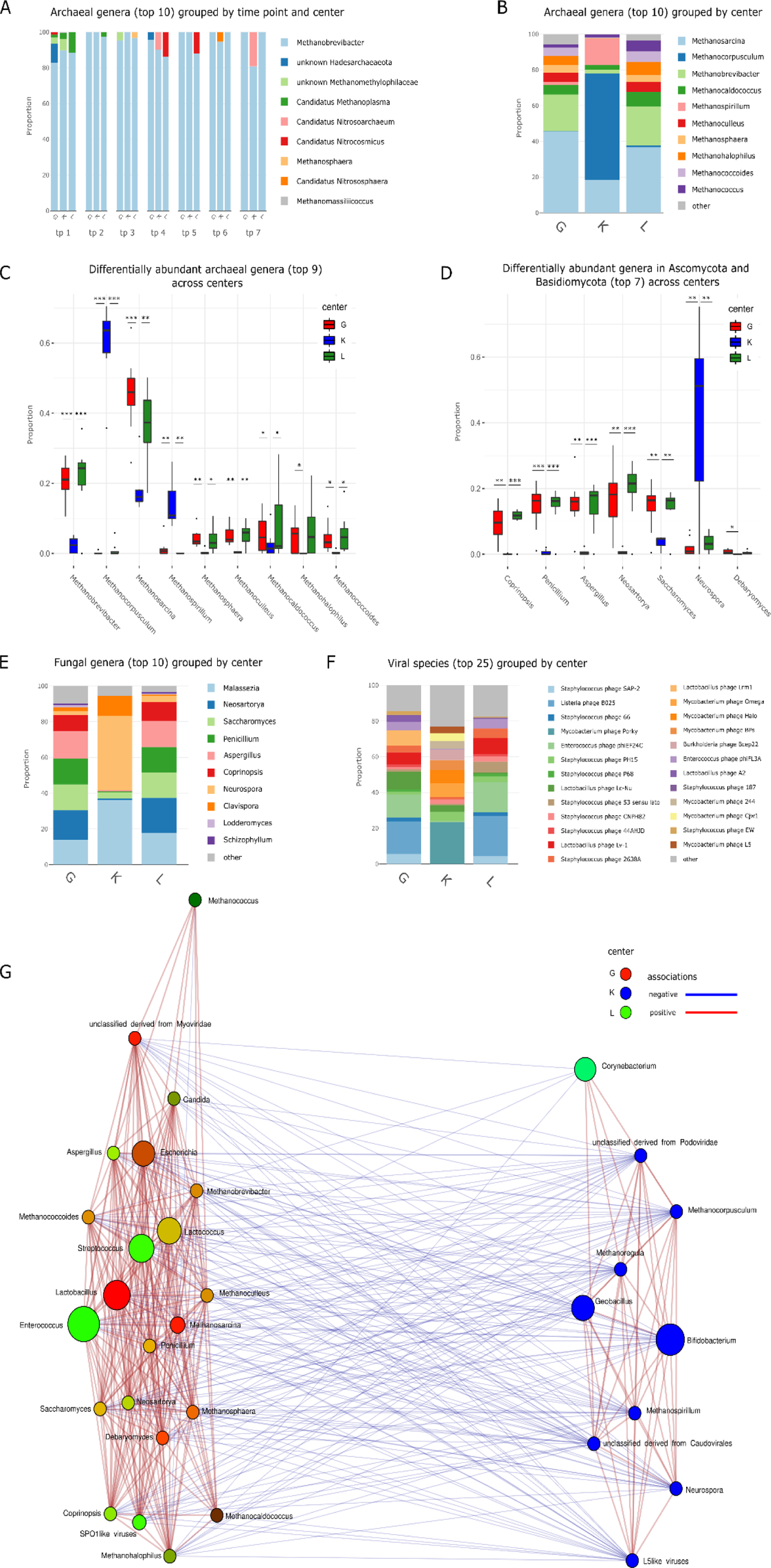
Overall distribution and abundances of microbial signatures of different domains, according to the different centres. (A) Stacked bar plot of relative abundances of the top ten archaeal genera in the amplicon NGS data set, displayed per centre at time points (tp) tp1–7. (B) Stacked bar plot of the top ten relative abundances of methanogenic archaeal genera in the MGS (metagenomic) dataset for time point tp7 per centre. (C) Box plot of relative abundances of the top nine methanogenic archaeal genera per centre. (D) Box plot of relative abundances of the top seven genera of Ascomycota and Basidiomycota per centre. (E) Stacked bar plot of the relative abundances of the top ten genera of Ascomycota and Basidiomycota in the MGS dataset for time point tp7 per centre. (F) Stacked bar plot of the top 25 relative abundances of phage species in the MGS dataset for time point tp7 per centre. (G) Network analyses of the ten most differentially abundant genera of each methanogens, ascomycota/basidiomycota, phages and bacteria; G in red, K in blue, L in green; significance levels are indicated with asterisks for *p* < 0.001 (***), *p* < 0.01 (**), *p* < 0.05 (*)

**Figure 2:**
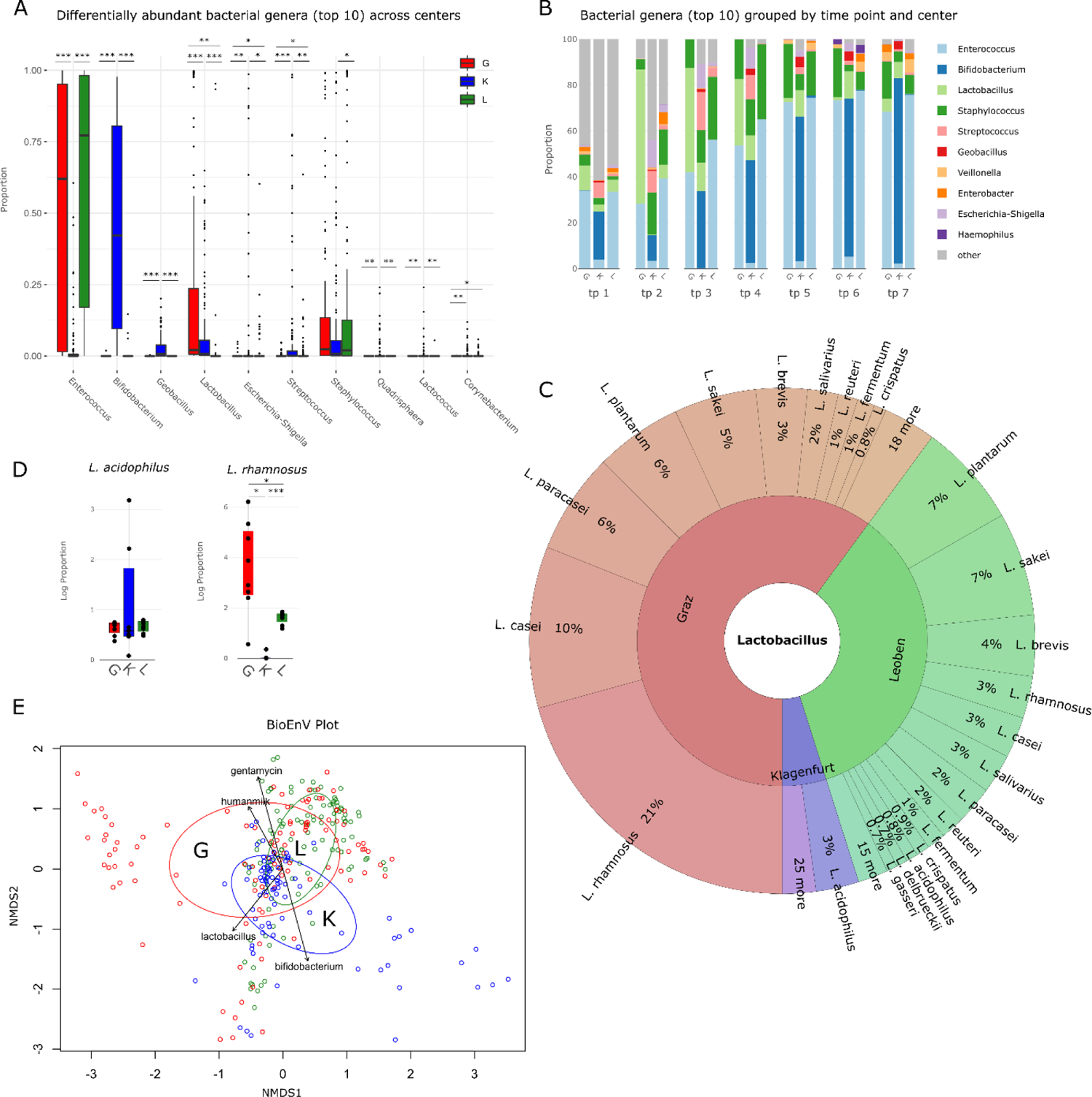
Distribution of bacterial taxa between the centres and influence of probiotic species. (A) Box plot of relative abundances of the top ten bacterial genera per centre; (B) Stacked bar plot of relative abundances of top ten bacterial genera in the amplicon NGS data set per centre each at timepoints tp1–7. (C) Krona chart of the distribution of species of the *Lactobacillus* genus between the centres. (D) Log percentages of relative abundance of probiotically administered *Lactobacillus* genera (i) *L. acidophilus* and (ii) *L. rhamnosus*. (E) Biplot of BioEnv with correlations of the Euclidean distances for the metadata of dissimilarities between the centres (administration of gentamicin, of probiotic *Lactobacillus* or *Bifidobacterium* and human milk). G in red, K in blue, L in green; significance levels are indicated with asterisks for *p* < 0.001 (***), *p* < 0.01 (**), *p* < 0.05 (*)

Unlike previous reports which concluded that infants generally do not carry archaea until three years of age, our archaea-focused approach enabled us to successfully detect 290 different ASVs with amplicon-based analyses and 75 different archaeal species with metagenomic-based sequencing. In particular, *Methanobrevibacter* was abundant, as it was detected in all infants in at least one sample and across all time points. In addition to typical human-associated archaea, *Methanobrevibacter*, *Methanosphaera*, Methanomethylophilaceae (incl. Methanomassiliicoccus) and various Nitrososphaeria (likely derived from skin sources (Moissl-Eichinger et al., 2017; Probst et al., 2013) (**Fig. 1A**), abundant signatures of *Methanosarcina* and *Methanocorpusculum* were additionally identified using shotgun metagenomics (**Fig. 1B**), indicating that even VLWB newborns are in contact with a wide diversity of archaea. Archaea reflected the centre, with, for example, *Methanocorpusculum* and *Methanospirillum* being significantly more abundant in samples from Klagenfurt (K) (*Methanospirillium*, *p* < 0.001, *p* < 0.01), than in Graz (G) and Leoben (L) (**Fig. 1C**). *Methanobrevibacter* and *Methanosarcina*, in contrast, were significantly more abundant in G and L than in K (Welch’s *t*-test, *p* < 0.01).

The contribution of fungal signatures to the overall microbiome was largely limited (also probably due to the application of antifungals) to Basidiomycota and Ascomycota (**Fig. 1D, E**), which, however, also revealed a centre-specific pattern: *Neosarorya*, *Penicillum*, *Aspergillus* and *Coprinopsis* (increased in G and L) were antiparallel to *Malassezia*, *Neurospora* and *Clavispora*, which were increased in K.

In order to confirm the centre-/regimen-specific profiles of the multiple-component microbiome data (further details, see below), a network analysis was performed based on the ten most differentially abundant genera of bacteria, methanogens, ascomycota/basidiomycota and phages (**Fig. 1G**). Taxa assigned exclusively to K formed a separate cluster and showed negative associations with taxa assigned to G and L, with *Corynebacterium* being an exception. Taxa assigned to G and L showed overall positive associations and clustered together. Some taxa were even shared by both centres (e.g. *Methanococcus*, *Candida*), suggesting an interaction between the domains.

The clustering of G and L bacterial, archaeal, fungal and phagal microbiomes in the network analyses was somewhat unexpected, but indicated that extrinsic factors might have this strong separating effect, including the HM feeding and gentamicin antibiotic prophylaxis in G and L, and the *Bifidobacterium*-based probiotics application in K.

### Supplemented *Bifidobacterium* suppresses natural pathobiont colonizers and co-administered lactobacilli

The bacteriomes of the infants were mainly characterized by the predominance and differential abundance of six bacterial key taxa, namely *Enterococcus, Bifidobacterium, Lactobacillus, Staphylococcus, Geobacillus* and *Escherichia* (**Fig. 2A**).

*Enterococcus* was found to predominate the bacterial microbiome in G and L (77%), together with less abundant *Lactobacillus* and *Staphylococcus*, the relative abundance of which decreased with maturation. In contrast, the K samples were dominated by *Bifidobacterium* at each time point and reached a relative abundance of > 82% at tp7 (**Fig. 2B**). Thus, G and L were dominated by a typical colonizer of the GI in preterm infants, whereas K samples showed an overall predominance of a supplemented probiotic taxon. The phenomenon of detection of *Geobacillus* signatures exclusively in K samples has been discussed previously (Kurath-Koller et al., 2020) and in the following chapter.

Notably, 16 of all 89 phage species were strongly associated with bacterial key species, resulting in a centre-specific, strongly differing phage profile (**Fig. 1F**). No bacteriophages were identified for *Bifidobacterium*, *Escherichia-Shigella* and *Geobacillus*; however, other phages from key species correlated with the relative abundance of their host, exemplified by *Streptococcus* in **Suppl. Fig. 1**. In particular, phages targeting *Lactobacillus* (Welch’s *t*-test; G:K *p* = 0.00289, K:L *p* = 0.00606) and *Enterococcus* (Welch’s *t*-test; G:K *p* = 0.0011, K:L *p* = 0.00612) were lowest in K.

Shotgun metagenomics confirmed the significantly differential abundance of *Bifidobacterium* in the three centers (K 82%; G 0.07%; L 0.08%; Welch’s *t*-test, K:G *p* < 0.001, K:L *p* < 0.001, G:L *p* > 0.05). Although *B. longum* subsp. *infantis* was the only *Bifidobacterium* administered in K, nine additional *Bifidobacterium* species were detected, with six species present in all K infants (*B. longum*, *B. dentium*, *B. breve*, *B. bifidum*, *B. animalis*, *B. adolescentis*). *B. longum* accounted for 95% of all reads from *Bifidobacterium*, and indeed this taxon was verified as *Bifidobacterium longum* subsp. *infantis* by read-mapping against six *Bifidobacterium* genomes known and isolated from human (infant) faeces (see Material and Methods and **Suppl. Table 2**). These analysis results confirmed that the major *Bifidobacterium* signatures in K infants were indeed from the administered probiotic. Of note, the signatures of *B. longum* subsp. *infantis* are abbreviated as *B. infantis* in the following.

Naturally, bifidobacteria are uncommon in the premature infants’ GI in the first days of life and start colonizing naturally beginning from week four and on (Alcon-Giner et al., 2020; Underwood and Sohn, 2017). Our data also indicate an absence of bifidobacteria in preterm infants who did not receive it via supplementation. Furthermore, administration of *B. infantis* resulted in combative, effective suppression of other bacteria, including the co-administered *Lactobacillus*: Although both probiotic species, *B. infantis* and *L. acidophilus*, were administered in equal amounts in K, *L. acidophilus* was initially suppressed by the predominant *Bifidobacterium*, probably due to its different metabolic capacities (0.21% relative abundance of *L. acidophilus* at tp7 vs. 75.69% relative abundance of *B. longum*).

### Probiotic administration of lactobacilli increases their natural abundance and diversity

In total, 27 species of the genus *Lactobacillus* (see details on classification issues in Star Methods), were detected by shotgun sequencing with different profiles between the centres (**Fig. 2C**). Probiotic lactobacilli were administered only in K (*L. acidophilus*) and G (*L. rhamnosus*). The presence of these lactobacilli in the infants’ intestinal samples was confirmed by amplicon sequencing but also by read-mapping (metagenomic dataset) against reference genomes of *L. rhamnosus* (UMB0004) and *L. acidophilus* (LA-14) (**Fig. 2D**).

*Lactobacillus rhamnosus* was also detected with low relative abundance (3%) in L, where it was not administered, suggesting that this species is indeed part of the natural infant gut microbiome of preterm infants and is likely transmitted through breastfeeding (Łubiech and Twarużek, 2020). In G, *L. rhamnosus* exhibited the highest abundance in G infants (21%) indicating a seven-fold increase in probiotic lactobacilli through its probiotic administration. K had the lowest absolute *Lactobacillus* abundance, with 56% of all *Lactobacillus* reads representing *L. acidophilus*, the species administered (K: 45,268 reads; G: 10,207; L: 12,431). Similar to *L. rhamnosus*, *L. acidophilus* was also detected in almost all infants in all centres, even when not supplemented as probiotics (**Fig. 2D**).

Although *L. acidophilus* was administered in K, the competitive growth of *B. infantis* administered at the same concentration seems to have efficiently suppressed/displaced the *Lactobacillus* species, as reflected by an *L. acidophilus*/ *B. infantis* ratio of ∼1:300 and *Lactobacillus*/ *Bifidobacterium* ratio of ∼1:200 (for genera see **Suppl. Table 3**).

The dominance of *Bifidobacterium* over *Lactobacillus* could also be underlined with the BioEnv Biplot (**Fig. 2E**). This plot shows the metadata whose Euclidean distances have the maximum (rank) correlation with community dissimilarities. In particular, the administration of *Bifidobacterium* and gentamicin correlated strongly with dissimilarities between the centres, whereas the administration of lactobacilli and HM correlates to a lesser degree.

### Key species are reflected by MAGs and active replication of probiotic species could be inferred

Samples from tp3 (days 5–8) and tp7 (days 13–21) allowed for deep metagenomic sequencing and genome binning. MAGs were obtained for *B. infantis* (K), *G. stearothermophilus* (K), *E. faecium* (G, L), *E. coli* (K), *L. rhamnosus* (G), *Veillonella parvula* (G, inactive), *Klebsiella oxytoca* (G, inactive) and *Escherichia flexneri* (L, inactive). Successful binning of the bacterial genomes followed the scheme of probiotic supplementation, with *B. infantis* MAGs in K samples and *L. rhamnosus* MAGs in G samples. Of the L samples, only MAGs of *Enterococcus faecium* were obtained (**Fig. 3A**). Application of iRep (Brown et al., 2017) revealed that MAGs corresponded to actively replicating bacteria (iRep values >1), indicating successful niche colonization by probiotic (*B. infantis* and *L. rhamnosus*) or naturally predominant bacteria (*E. faecium*) (**Fig. 3A**).

**Figure 3:**
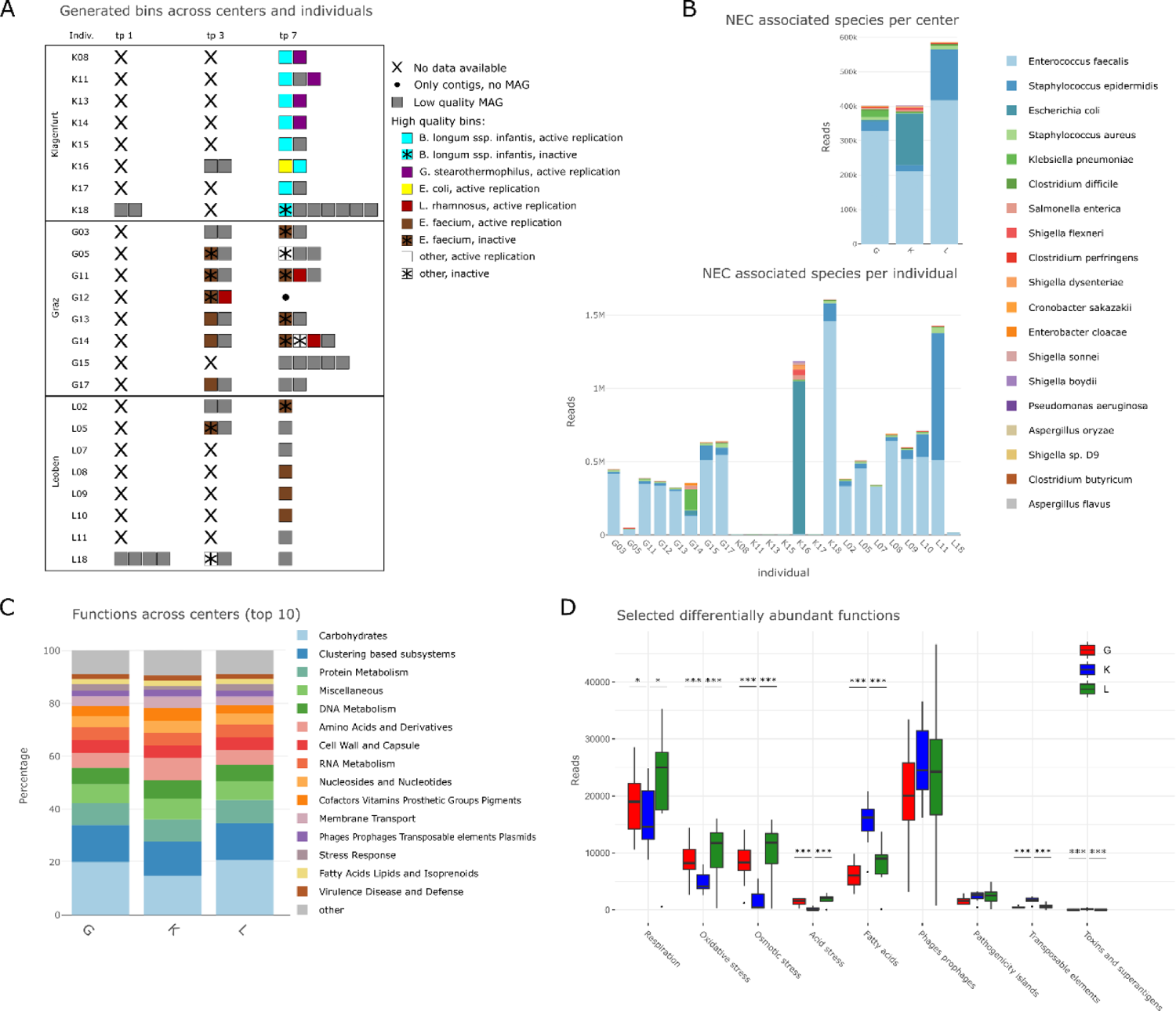
Replication values for MAGs, distribution of potentially NEC causing microbes, and specific functions. A) Retrieved MAGs/contigs per centre, individual and time points tp1, tp3 and tp7: availability, quality, iRep replication status [min: 1.348, max: 1.998, mean: 1.654] and taxonomic placement; B) Read numbers of microbial species that were correlated with NEC previously, per centre and per individual; C) top ten microbiome functions per centre; D) reads of ten selected differentially abundant functions per centre. G in red, K in blue, L in green; significance levels are indicated with asterisks for *p* < 0.001 (***), *p* < 0.01 (**), *p* < 0.05 (*).

Notably, high-quality *G. stearothermophilus* genomes with iRep values above 1.41 were isolated from four out of eight infant samples from K (tp7) (see also (Kurath-Koller et al., 2020)). *G. stearothermophilus* is a frequent, most probably harmless contaminant in milk plants (Kumar et al., 2021), which probably transforms to an active form during formula milk preparation and is then ingested by the infant. Thermophilic *G. stearothermophilus* grows in the temperature range of 40–70°C, with optimal growth rates achieved at 55–65°C (Durand et al., 2015) (Nazina et al., 2001); thus, an active proliferation in the infant GI is unlikely. *Staphylococcus* MAGs could not be retrieved, despite its high abundance and identification as a key microorganism in this study (**Fig. 3A**).

### Low level occurrence of diverse types of potentially pathogenic bacteria

Next, we searched specifically for signatures that had been associated with outbreaks or cases of NEC in previous reports (Underwood and Sohn, 2017). In our shotgun metagenomic dataset, we identified *E. faecalis* as having the highest abundance in this dataset, while *S. epidermidis*, *E. coli* and others were found in varying amounts in samples from tp7 (**Fig. 3Bi**). While the infants from the other centres showed a more or less homogenous presence of potential NEC-causing microorganisms, the hits in K samples concentrated solely on two infants, K16 and K18 (**Fig. 3Bii**). Reads of *E. faecalis* derived mainly from K18, whereas a high number of *E. coli* signatures originated almost exclusively from K16, the only infant in this subset that developed NEC later on. Furthermore, a MAG of probably active *E. coli* could be obtained from this infant at tp7 (i.e. 14 days after birth), suggesting the possible initial bloom of *E. coli* already at this early time point before NEC is usually diagnosed. As *E. coli* MAGs were not retrieved from any other sample, our data support the potential for microbiome analyses to be used for NEC monitoring and diagnosis (**Fig. 3A**).

### Functional profiles mirror earlier gut maturity in *Bifidobacterium*-supplemented infants

We also profiled microbiome functional characteristics at tp7 given the sufficient sequencing coverage obtained at this time point (**Fig. 3C**). K samples showed an overall significantly reduced level of genes involved in oxidative stress, osmotic stress, acid stress and respiration (Welch’s *t*-test, *p* < 0.01), indicating early maturation of the infant gut and successful establishment of anaerobic conditions. Indeed, *Bifidobacterium* is an anaerobic bacterium, whereas lactobacilli are considered facultative anaerobes. The early maturation of the GI in K infants was further confirmed by the increased relative abundance of genes involved in polysaccharide metabolism, allowing the degradation of more complex sugars, which was also confirmed by metabolomics (**Fig. 3D**; see also below).

Notably, the number of genes related to toxins and superantigens, as well as of transposable elements (involved in the distribution of pathogenic genomic features), were significantly higher in K samples (Welch’s *t*-test, toxins and superantigens *p* < 0.01; transposable elements *p* < 0.001), suggesting a potentially higher bacterial pathogenesis signature in these infants.

Next, we performed NMR-based metabolomics to determine the concentration of short-chain fatty acids (SCFAs) and complex sugars in the infants’ stool samples. We found a general increase in acetic acid, formic acid, valeric acid and butyric acid over time in all centres, regardless of the microbiome composition or probiotic supplementation (**Suppl. Fig. 2A-D**). However, unlike previous reports, we observed an unexpected spike in propionic acid at tp3 in all centres (**Suppl. Fig. 2E**) (Wang et al., 2021). We hypothesize that this spike in propionic acid is related to a delayed uptake of propionate by the intestinal epithelium during maturation.

### Human milk supports *Bifidobacterium* by associated HMO conversion which is impaired by formula milk feeding

Genes involved in carbohydrate metabolism were the most abundant functional features in our shotgun metagenomic dataset. Notably, the samples from K had a significantly lower proportion than the other centres (Welch’s *t*-test, K:G *p* = 0.006, K:L *p* = 3.24e-5). Upon further examination, particularly genes involved in the metabolism of monosaccharides were significantly lower in K than in G and L (Welch’s *t*-test, G:K *p* = 1.56e-3, K:L *p* = 7.88e-5) (**Fig. 4A**). The significantly lower availability of monosaccharides in K was confirmed by a representative, metabolomic-based quantitative assessment of D-glucose and D-fructose. However, an overall increase of both compounds was observed in all centres over time (**Suppl. Fig. 2F,G**). Similarly, genes involved in metabolism of di- and oligosaccharides were substantially reduced in K samples (Welch’s *t*-test, K:L *p* = 0.0018; K:G *p* = 0.093; **Fig. 4A**). In contrast, the gene proportion involved in polysaccharide metabolism was found to be significantly increased in K samples (Welch’s *t*-test, K:G *p* = 2.50e-5, K:L *p* = 5.10e-5) (**Fig. 4A**), indicating a higher genetic potential for the metabolism of complex sugars in K.

**Figure 4:**
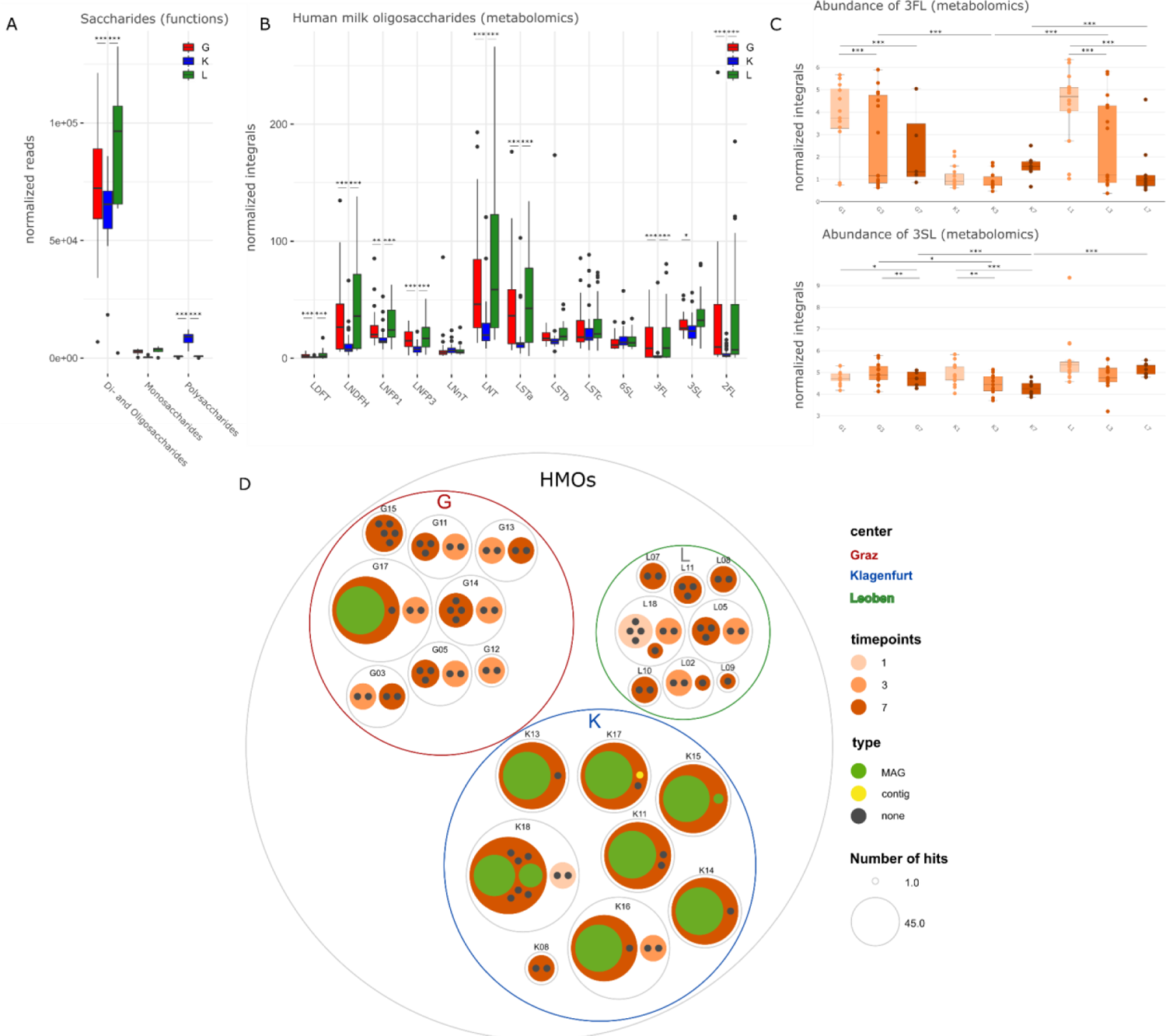
Distribution patterns of sugars and HMOs. A) Differentially abundant functions (metagenomic dataset) associated with saccharides (di- and oligosaccharides, monosaccharides and polysaccharides) between the centres; B) differentially abundant HMOs between centres (metabolomic dataset); C) differential abundance of the HMOs i) 3’-fucosyllactose 3’FL and ii) 3’-sialyllactose 3’SL between centres at tp1, tp3 and tp7; D) circle packing plot displaying the numbers of hits for HMO gene clusters found in MAGs and contigs at the different time points. Each circle represents one infant. Colours of the dots indicate where the hit occurred, on MAGs (green) or if it could be only assigned to a contig (yellow) or none (grey); the size of the dots indicate the number of hits. G in red, K in blue, L in green; significance levels are indicated with asterisks for *p* < 0.001 (***), *p* < 0.01 (**), *p* < 0.05 (*)

To answer the question regarding the genetic potential for complex HMO degradation, we searched for HMO gene clusters in the obtained MAGs and contigs. Indeed, the potential for HMO metabolism was notably higher in samples from K (**Fig. 5D**). The total hits with HMO gene clusters were G: 45, K:307, L:0, indicating a seven-fold higher numbers of HMO genes in K than in G. Moreover, only one MAG from G17 possessed the potential to convert and digest HMOs, in contrast to all infants from K with at least one MAG (**Fig. 5D**).

**Figure 5:**
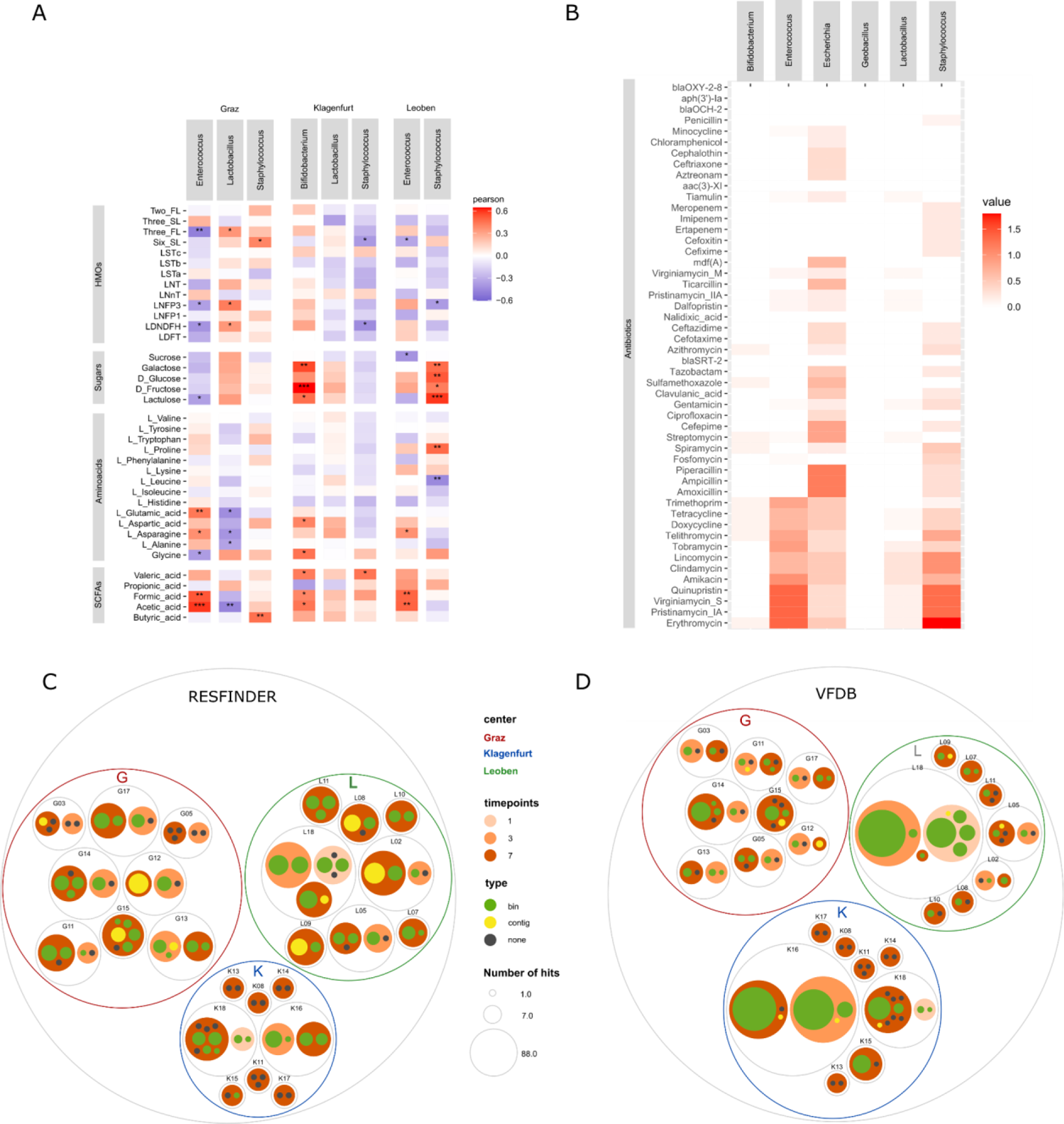
Results of taxonomic correlation analyses and distribution patterns of metabolites, antibiotic resistances and virulence facts. A) Metabolites measured with NMR correlated with bacterial key genera in the three centres; metabolites of the groups of human milk oligosaccharides (HMOs), sugars, amino acids and short-chain fatty acids (SCFAs); B) correlation of antibiotic resistance genes with bacterial key genera. Circle packing plot of C) resistance genes and D) virulence factors in the centres. Each circle represents an infant and is split into the three MGS sequenced time points. Colours of the dots indicate where the hit occurred, on MAGs (green) or if it could be only assigned to a contig (yellow) or none (grey); the size of the dots indicates the number of hits.

Using metabolomics, we assessed HMOs in the preterm stool samples. A total of 13 HMOs were detected at tp1, tp3 and tp7 (**Fig. 4B**). For the fucosylated HMOs (2’-fucosyllactose [2’FL], 3’fucosyllactose [3’FL], lacto-N-ducopentaoise I [LNFP1], lacto-N-fucopentaose III [LNFP3], lacto-N-difuco-hexaose [LNDFH], lactodifucotetraose [LDFT]) as well as LS-tetrasaccharide a [LSTa] and lacto-N-tetraose [LNT], a significantly decreased amount for K samples was detected as compared to G and L (Welch’s *t*-test, *p* < 0.05) (**Fig. 4Ci**). For syalisated HMOs (such as 3’-sialyllactose [3’SL], 6’-sialyllactose [6’SL], LS-tetrasaccharide b [LSTb], LS-tetrasaccharide c [LSTc], but also lacto-N-*neo*tetraose [LNnT]), few differences between centres or time points were observed (**Fig. 4Cii**). We conclude that the K microbiomes have the largest potential to degrade HMOs effectively; however, this process is impaired by the lowered availability of HMOs by preferred formula feeding.

### Regimens and their key taxa correlate with crucial metabolites, antibiotic resistance genes and virulence factors

We were interested in the correlations among the six microbial key players with HMOs, carbohydrates, amino acids and short-chain fatty acids, as measured using metabolomics (**Fig. 5A**) (additional metabolites are shown in **Suppl. Fig. 3**). *Bifidobacterium* (K) showed positive correlations with several carbohydrates (galactose and fructose), and weaker ones, with certain amino acids and short-chain fatty acids. It appears under HMO-depleted conditions, *Bifidobacterium* uses galactose and fructose as alternative substrates for its carbohydrate metabolism, producing formate and acetate. In the absence of *Bifidobacterium* (G and L), the role of formate and acetate production is taken over by *Enterococcus*, indicating its importance for early SCFA production, probably from citrate or pyruvate (Ramsey et al., 2014). *Staphylococcus* showed an inconsistent pattern across the centres, supporting our hypothesis that this microorganism played a smaller role for the preterm GIT. The metabolic pattern of *Lactobacillus* (G, K) could not be clearly resolved, as lactate was not included in the metabolomic approach.

The antibiotic gentamicin (used in G and L; Table 1) has a broad spectrum of activity including: *Enterobacter, Escherichia, Proteus, Klebsiella, Pseudomonas, Serratia* and *Staphylococcus* (Corporation, 2012). Surprisingly, the influence of gentamicin on these genera was not evident in the preterm microbiome samples, and only limited perturbations were observed. However, it should be noted that, in this study, gentamicin was administered enterally and not intravenously, which may result in an altered mode of action. Acquired resistance to the gentamicin administered was rarely detected.

Overall, clear differences were observed in the antibiotic resistance (AMR) profiles between the key genera (**Fig. 5B**) and centres (**Fig. 5C**). AMR potential of the MAGs was substantially reduced in K samples as compared to samples from G and L (G: 93 hits, K:35 hits, L:123 hits), once again highlighting the influence of *Bifidobacterium* on microbiome composition and function. Of the AMR signatures detected, most were positively correlated with the potential NEC/sepsis pathogens *Enterococcus*, *Staphylococcus* and *Escherichia,* including resistance against β-lactams and erythromycin (**Fig. 5B**). In contrast, AMR profiles were highly limited in the probiotic genera *Bifidobacterium* and *Lactobacillus*, underscoring the safety of probiotic administration. *Geobacillus* was also found to not encode AMR genes; therefore, it did not appear to pose an increased health risk.

The virulence factor analysis results show that K infants had fewer pathogenic factors in their microbiomes than G and L infants (G:64 hits, K: 193 hits, L:173 hits) (**Fig. 5D**). Enrichment in *E. coli*-associated virulence traits was also observed in two infants in K and L (K16 and L18). Only six out of 431 hits were observed for *Staphylococcus* in six infants. We can assume that *Staphylococcus* is a key player in the context of its abundance, but not in the context of either replication or virulence factors. As expected, no virulence factors were found for the other key players, including *Bifidobacterium*, *Lactobacillus* and *Geobacillus*.

## Discussion

NEC prophylaxis and therapy have become a central aspect in the clinical management of VLBW preterm infants. Although probiotics, human milk and antibiotics are used by many NICUs to prevent NEC, to our knowledge, an in-depth, systematic comparison of different preventive regimens had not yet been performed. In this study, we show that NEC prophylaxis not only impacts the bacterial gut microbiome composition and function strongly, but also drives strong centre-specific patterns observed in the fungal, archaeal and viral parts of the microbiome. This is particularly important with respect to archaea and phages/viruses, as NEC prophylaxes are not directly administered to change this part of the microbiome.

Our findings show that *Bifidobacterium* supplementation (as compared with other probiotics, nutritional and antibiotic treatments) appears to have the most beneficial effects on the early gastrointestinal microbiome, as reflected by centre K having the lowest NEC rates. Our iRep analyses and the steady increase in relative abundance (**Fig. 2B**) strongly suggest that *Bifidobacterium* actively replicates and thus successfully colonizes the GIT during the administration period. This is supported by the presence of HMO gene clusters and low levels of HMOs in the stool. Although *Bifidobacterium* does not yet appear to naturally colonize the GIT of preterm infants in large numbers, the reliable health benefits of supplemented *Bifidobacterium* have been demonstrated in multiple ways (Picard et al., 2005), including the negative correlation between *Bifidobacterium* and opportunistic pathogens (Klopp et al., 2022).

Unlike *Bifidobacterium*, a variety of lactobacilli, including *L. acidophilus* (administered in centre K) and especially *L. rhamnosus* (administered in G), have been observed as natural colonizers of the GIT of preterm infants. In this study, we consider human milk (HM) to be the most likely source, as HM naturally contains large amounts of live lactobacilli (Lyons et al., 2020). For example, *L. rhamnosus* was successfully isolated from 8.13% of all human milk samples examined by Lubiech et al. (Łubiech and Twarużek, 2020) and was also detected in culture-independent assays alongside other, more frequently occurring *Lactobacillus* species (*L. salivarius, L. fermenting, L. gasseri* (Łubiech and Twarużek, 2020; Ruiz et al., 2019; Soto et al., 2014). It should be noted, however, that the HM was pasteurized, especially if it came from a milk bank, and pasteurization may obviously reduce the chance that live lactobacilli are transmitted. As all except one infant in the metagenomic subset were born via C-section and not vaginally, vertical transfer from the maternal vaginal microbiome is untenable in this case.

The dynamics of naturally occurring and supplemented (non-natural) probiotic strains are interesting to study, as supplemented bacteria have to “invade” an ecosystem that is evolving to provide colonization resistance against other, potentially pathogenic bacteria. To avoid disrupting this process, and also considering that naturally occurring bacterial residents are likely to persist over a longer period of time (Dalby and Hall, 2021), an ecologically oriented probiotic choice would tend to include supplementation with *L. rhamnosus* or another beneficial preterm *Lactobacillus* strain.

However, we and others observed that the co-administration of *Bifidobacterium* and *Lactobacillus* in equal amounts results in the overgrowth and predominance of *Bifidobacterium* over *Lactobacillus* (Abdulkadir et al., 2016; Alcon-Giner et al., 2020). The absence of replicating *Lactobacillus* MAGs in the presence of replicating *Bifidobacterium* MAGs leads us to hypothesize that *Lactobacillus acidophilus* cannot successfully colonize the GIT of preterm infants when *Bifidobacterium longum* subspecies *infantis* is co-administered. On the other hand, it may be that *Lactobacillus* plays a pioneering role in anaerobic bifidobacterial colonization by removing oxygen from the GIT (Koskella et al., 2017). This supporting role of *Lactobacillus* tends to underscore the benefits of taking a multispecies probiotic approach.

Most importantly, administration of *Lactobacillus* alone had no appreciable effect on the composition or function of the gut microbiome and did not result in a substantial increase in *Lactobacillus* colonization as seen before (Martí et al., 2021). Potentially pathogenic bacteria and antibiotic resistance genes were also not substantially reduced when *Lactobacillus* was administered alone.

We argue that the GIT of VLBW infants per se is not a “normal” natural habitat, and the action of *Lactobacillus* may be too mild or too slow to rapidly support the establishment of a healthy microbiome. In contrast, *Bifidobacterium*, although not naturally occurring, proved to be a strong, stable and reliable keystone microbe of the nearly empty niche of the preterm GIT, efficiently suppressing pathogenic threats and microorganisms carrying antibiotic resistance genes.

Moreover, *Bifidobacterium* together with *Bacteroides* has been described as an effective converter of HMOs (Marcobal and Sonnenburg, 2012; Marcobal et al., 2010), producing substantial amounts of beneficial SCFAs (Esaiassen et al., 2018). The efficient conversion of HMOs and thus SCFA production is an extremely important process for premature infants, which is underscored by the finding that concentrations of HMOs in HM are substantially increased when the infant is born prematurely (Austin et al., 2019; Łubiech and Twarużek, 2020). In fact, the bifidobacteria found in the faeces of K infants exhibited a marked genetic ability to convert HMOs. This could be related to changes in the GIT environment such as lower pH or improved colonization resistance.

This capacity, however, could not be observed in our metabolomic analyses because the K infants were fed with (HMO-lacking) FM. In contrast, in centres L and G, where HMOs were administered in the natural form of HM, the small natural proportion of microbial HMO converters was too low for observable efficient turnover. Consequently, only the simultaneous administration of HMOs <u>and</u> HMO-converters would result in optimal utilization of the health benefits for the infants. This highlights the importance of combining the right probiotic with the right diet.

Furthermore, clinically relevant findings from our study are related to enteral antibiotic administration, as gentamicin was administered in two NICUs, G and L. Indeed, the prophylactic enteral, but not parenteral, administration of antibiotics has been shown to substantially decrease NEC rates (Birck et al., 2016; Bury and Tudehope, 2001), which is also reflected in the low number of NEC cases observed in centre G (Schmolzer et al., 2006). To date, no single causative microbial agent of NEC has been described; thus, the antibiotics used, such as gentamicin, must cover a broad spectrum. Our study was not designed to investigate the performance and efficiency of gentamicin in eliminating specific bacteria. However, we did not find consistent negative correlations between gentamicin administration with certain taxa or the occurrence of gentamicin resistances in the microbiome at tp7. Nevertheless, we cannot rule out a negative effect on concomitantly administered probiotic bacteria.

In several studies, intravenous antibiotic administration at a young age has been associated with adverse health outcomes later in life (Alexander et al., 2011; Fan et al., 2017; Michael Cotten et al., 2009). It is also proposed that antibiotic prophylaxis does not reduce NEC incidences but may rather increase the risk for high-risk premature infants of NEC (Jin et al., 2019). Nevertheless, in these studies antibiotics were administered enteral, not intravenously, which needs to be evaluated strictly differently. Still, the prophylactic use of antibiotics must always be weighed against the potential risk: On the one hand, prophylactic administration of antibiotics probably minimizes the outbreak of pathogenic bacteria, especially since infections in premature infants develop alarmingly rapidly, and the success of treatment is time-critical. On the other hand, antibiotics could also suppress the growth of beneficial (probiotic) bacteria, which is also underlined by our study results showing that probiotic *Lactobacillus* and *Bifidobacterium* carry few AMR genes and are therefore more susceptible for antibiotics. The long-term effects of antibiotic administration at such an early, vulnerable age are difficult to predict. In general, AMR is a global threat (World Health Organization, 2015) and their horizontal gene transfer to pathogenic bacteria might also be implicated in NEC.

Our study has several strengths and limitations. Overall, due to the extensive analysis performed, the study cohort was kept rather small, and the survey period was limited to the first weeks of life. Unfortunately, the early (meconium) samples could not be used for metagenomic analyses because of their exceptionally low microbial biomass, so we had to focus on tp7 for detailed functional assessments. However, we successfully and comprehensively conducted a multicentre study in which we analysed the longitudinal composition of the microbiome of 54 VLBW preterm infants using amplicon-based and metagenomic sequencing, which also enabled us to elucidate the contribution of archaea, fungi and phages. A large wealth of taxonomic and functional data were obtained, and analyses revealed the HMO conversion potential and the emergence of antibiotic resistance. In addition, genetically detected functions could be effectively combined with well-found metabolomic analyses.

## Conclusion

Our study provides a solid basis for further evaluation and analyses. We found that the combination of feeding HM and administering Bifidobacterium during the first weeks of life in VLBW infants could be a promising synergistic approach. Overall, all treatment regimens analysed in this study resulted in NEC rates well below the global average, confirming the excellent and strategic management of this devastating disease in our NICUs. These therapies, or perhaps a novel combination thereof, could serve as a model for implementing similar strategies to combat NEC in other NICUs worldwide.

## Data Availability

All data produced are available online at

https://github.com/CharlotteJNeumann/preterm_shared

## Acknowledgements

We highly appreciate the contributions of Raimund Kraschl, Claudia Kanduth, and Barbara Hopfer. This research was funded in in part by the Austrian Science Fund (FWF) [DOC 31 DP-iDP and P32697, given to CME]. For the purpose of open access, the author has applied a CC BY public copyright licence to any author-accepted manuscript version arising from this submission. TM was supported by Austrian Science Fund (FWF) grants P28854, I3792 and DK-MCD W1226, the Austrian Research Promotion Agency (FFG) grants 864690 and 870454; the Integrative Metabolism Research Center Graz; Austrian Infrastructure Program 2016/2017, the Styrian Government (*Zukunftsfonds*) and BioTechMed-Graz (Flagship project DYNIMO). BR was supported in part by a grant of the parent association “*Kleine Helden –Initiative für Früh-und Neugeborene*”, Graz, Austria. LJH is supported by Wellcome Trust Investigator Awards (100974/C/13/Z and 220876/Z/20/Z); the Biotechnology and Biological Sciences Research Council (BBSRC), Institute Strategic Programme Gut Microbes and Health (BB/R012490/1), and its constituent projects BBS/E/F/000PR10353 and BBS/E/F/000PR10356. The authors acknowledge computational resources of the MedBioNode at the Medical University of Graz and the support of the ZMF Galaxy Team: Core Facility Computational Bioanalytics, Medical University of Graz, funded by the Austrian Federal Ministry of Education, Science and Research *Hochschulraum-Strukturmittel* 2016 grant as part of BioTechMed Graz.

The funding body had no influence on the study, collection, analysis and interpretation of data or on the manuscript content.

Conceptualization: BU, BR, SKK, CME Methodology: CJN, SKK, AM, CK, RK, MD, TM

Formal Analysis: CJN, AM, CK, RK, MD, MK, TM Investigation: CJN, CME

Writing - Original draft: CJN, CME Writing - Review & Editing: all authors Visualization: CJN

Supervision: CME, BR, BU, LH Project Administration: CJN, BU, CME

Funding Acquisition: BR, BU, TM, CME

## 11) Declaration of interests

The authors declare no competing interests.

## 12) Inclusion and diversity statement

We worked to ensure sex balance in the selection of participants, as well as authors. We further actively worked to promote gender balance in our reference list.

## Methods

### Study design

We conducted a prospective, triple-center cohort pilot study investigating the gut microbiome of preterm infants with a birthweight < 1,500 g in three Austrian neonatal intensive care units (NICUs). These centers (Klagenfurt, K; Leoben, L; Graz, G) used different regimens for NEC prophylaxis which are summarized in Table 1 and have been described in detail previously (Kurath-Koller et al., 2017) (Kurath-Koller et al., 2020). A detailed description of the study design is available in (Kurath-Koller et al., 2017) and first results have already been published elsewhere (Kurath-Koller et al., 2020).

In G, prophylaxis consisted of administration of probiotic *Lactobacillus rhamnosus* twice a day, nystatin, and enteral gentamicin. Probiotic bacterial species were also administered in K, namely *Bifidobacterium longum* subsp*. infantis* and *Lactobacillus acidophilus* in combination with fluconazole. In center L, no probiotic species, but enteral gentamicin and nystatin were used. Next to medication and probiotic supplementation, the feeding regimen also differed between the centers. In G and L, mainly human milk (HM) was provided. In K, enteral nutrition consisted mainly of formula milk (FM).

Between October 2015 and March 2017, stool samples were collected from preterm infants at those three centers. Inclusion criteria were birth weight < 1,500 g and survival in the first three weeks of life. Clinical data on the infants have been published recently (Kurath-Koller et al., 2020), with no significant differences between the centers (APGAR, sex, gestational age, gestational weight), except length of hospital stay (G: 72 K: 68.5 L:58; p=0.04)). In case of genetic diseases, syndromes or congenital anomalies or meconium ileus, infants were excluded from the study. A total of 54 infants were included in the study. The infants’ stool samples were collected every other day from meconium for the first two weeks of life, with each infant providing stool samples at seven time points (timepoints of samples were slightly variable (**see Suppl. Table 1**) due to the varying availability of fecal samples; average sampling time points were tp1: within day 1-3; tp2: day 4; tp3: day 6; tp4: day 8; tp5: day 10; tp6: day12; tp7: day 15). A total of 383 samples and 16 negative controls were collected.

The diagnostic criteria for NEC definition were the same in all three centers and followed the AWMF guideline with Bell criteria with modifications of Walsh. NEC incidence rates are 2.2% in K, 2.7% in G, and 4.6% in L (Kurath-Koller et al., 2020). The study received ethical approval from the local ethic committees (number 27-366 ex14/15) and written informed consent was obtained from the parents of the infants.

### Sample processing

#### DNA extraction, sequencing and metabolomics

Samples were processed as described in detail earlier (Kurath-Koller et al., 2020). Shortly, genomic DNA was isolated according to manufacturer’s instructions using the MagnaPure LC DNA Isolation Kit III (Roche). Targeted amplicon sequencing was performed for three different regions: one using universal primers but mainly targeting bacterial 16S rRNA gene sequences (515F/R926), the other aimed for optimized amplification of archaeal 16S rRNA gene sequences in a nested PCR (PCR1 244F/1041R, PCR 2 519F/806R) and the third of the ITS region of fungi (Turenne et al., 1999; White et al., 1990). See (Pausan et al., 2019) for detailed primer sequencing and PCR protocols. Sequencing was performed in paired-end run mode on an Illumina MiSeq with v3 600 chemistry and 300bp read length (Klymiuk et al., 2016). Raw reads are publicly available at the European Nucleotide Archive PRJEB37883. Raw reads were processed using Qiime2 v2019.1 to v2021.2 (Bolyen et al., 2019). Briefly, reads that were first quality filtered with DADA2 v2019.1.0 to v2021.2.0 (Callahan et al., 2016) were then denoised into Amplicon Sequence Variants (ASVs). The taxonomy was assigned with a Naive-Bayes classifier based on SILVA 132 for bacterial and archaeal signatures (Quast et al., 2013) (Bokulich et al., 2018). Potential contaminant ASVs were removed considering the sequenced negative controls with the R package decontam v3.9 in prevalence mode, is Contaminant setting and threshold 0.5 (Davis et al., 2018), (https://github.com/benjjneb/decontam/). Subsequently negative controls as well as signatures of chloroplasts and mitochondria were removed manually.

The genus *Lactobacillus* has recently been taxonomically re-structured (Zheng et al., 2020), in which the genus was divided into 25 separate genera. In this study, we continue to refer to the amended nomenclature and dimension of the genus *Lactobacillus*, as this work is a supplement to the previously published NGS data of this study, and we prefer to be consistent between those two publications. In addition, the taxonomic assignment of the datasets on which all other analyses are was performed using SILVA 132 (Quast et al., 2013) prior to the renaming event. In this publication, we mention only two representatives of the original *Lactobacillus* genus, namely *Lactobacillus rhamnosus* (now: *Lacticaseibacillus rhamnosus*) and *Lactobacillus acidophilus*, the latter remaining unchanged. It shall be noted, that for the important probiotic representatives of the amended *Lactobacillus* genus, the new names also begin with “L” and the abbreviations of the “*L.*” genus may continue to be used. (Zheng et al., 2020)

An initial insight into the bacteriome of the analyzed 54 infants, based on 16S rRNA gene amplicons was provided earlier (Kurath-Koller et al., 2020). In this subsequent data analysis, we intensify the analytical assessments and include shotgun metagenomics and functional metabolomics, to substantially deepen the understanding of the development within the first weeks. Further, we include information on the archaeal, fungal, and viral part of the gut microbiome as well as on their function.

We performed shotgun metagenomic sequencing of a subset of infants for three timepoints (tp1, tp3, tp7). Sequencing libraries were generated with the Nextera XT Library construction kit (Illumina, Eindhoven, the Netherlands) and sequenced on an Illumina HiSeq (Illumina, Eindhoven, the Netherlands; Macrogen, Seoul, South Korea). The raw reads were quality assessed with fastqc v0.11.8 (Andrews S, 2010) and filtered accordingly with trimmomatic v0.38 (Bolger et al., 2014) with a minimal length of 50 bp and a Phred quality score of 20 in a sliding window of 5 bp.

To map *Bifidobacterium infantis* reads against several *Bifidobacterium* reference genomes, quality filtered reads were mapped with bowtie2 v2.3.5 (Langmead and Salzberg, 2012) to reference genomes of *Bifidobacterium* isolated from humans (*B. longum* subsp. *longum* (JCM 1217^T^), *B. longum* subsp. *infantis* (JCM1222^T^), *B. longum* subsp. *suillum* (JCM 19995^T^), *B. catenulatum* subsp. *kashiwanohense* (JCM 15439^T^), *B. bifidum* (JCM 1255^T^) and *B. breve* (JCM 1192^T^); **Suppl. Table 2**) and mappable reads were kept with bedtools v2.29.2 (Quinlan and Hall, 2010) and samtools v1.9 (Li et al., 2009) from the quality-filtered fastq reads. This way reads classified as *B. infantis* could be validated as *Bifidobacterium longum* subsp. *infantis*.

Samples from time points tp1 and tp3 yielded only small amounts of DNA, so that library preparation or post-sequencing quality filtering failed for most samples. Thus, we focus mainly on data from tp7 for analysis and interpretation.

The obtained reads were analyzed both in a genome- as well as gene-centric way. For the gene-centric approach, data were annotated with diamond v0.9.25 (Buchfink et al., 2021) using blastx search against NCBInr database from 2019-07-19 and analyzed in the open-submission data platform MG-RAST according to the manual using default settings (Meyer et al., 2008), on taxonomic and functional (SEED subsystems (Overbeek et al., 2005)) level.

For the genome-centric analysis the reads were co-assembled with Megahit v1.1.3 (Li et al., 2015) by using the default setting “meta-sensitive” into contigs which were then binned with MaxBin2 v2.2.4 (Wu et al., 2014). After quality-checks of all bins with CheckM v1.1.0 (Parks et al., 2015) the bins were de-replicated with dRep v2.0.5 (Olm et al., 2017) to generate a list of representative metagenome assembled genomes (MAGs). Taxonomic classification of those MAGs was performed with GTDB-Tk v1.5.1. Replication rates were determined with iRep v1.1 (Brown et al., 2016). Indices for MAGs with the following default parameters were included: ≥ 75% completeness; ≤ 175 fragments / Mbp sequence; ≤ 2% contamination; ≥ 5 kbp scaffold length; min cov = 5; min wins = 0.98; min r^2 = 0.9; GC correction min r^2 = 0.0.

A subset of 123 stool samples for tp1, tp3 and tp7 (corresponding to metagenomic sequencing) were analyzed with untargeted NMR (nuclear magnetic resonance spectroscopy) for several metabolites in house as described previously (Kumpitsch et al., 2021). In short, methanol water was added to the samples, cells were lysed, lyophilized, and mixed with NMR buffer. NMR was performed on an AVANCETM Neo Bruker Ultrashield 600MHz spectrometer equipped with a TXI probe head at 310 K and processed as described elsewhere (Alkan et al., 2018).

### Data analysis, statistics and visualization

Network analysis was performed with the web tool Calypso (Zakrzewski et al., 2017) on amplicon and metagenomic sequencing taxonomy data. Microbial composition analysis and visualization as stacked bar charts were performed in R v4 (R-Core-Team, 2021) using the Microbiome Explorer package (Reeder et al., 2021), as was correlation analysis of abundance of specific bacterial genera with their phages. Differential abundance was plotted in R (R-Core-Team, 2021) using the ggplot2 package (Wickham, 2016). A pie chart for the *Lactobacillus* genus was created using Krona charts (Ondov et al., 2011). The BioEnv Biplot for bacterial dissimilarity of the groups was created using the vegan package (Oksanen et al., 2015) in R. Significance of differential abundance was calculated in STAMP using a two-sided Welch’s t-test with evaluating significance with p-values <0.05 (*), <0.01 (**) and <0.001 (***).

### Metabolite correlation with taxonomic information

Metabolites measured by NMR were then correlated with the relative abundance of genera and/or species of NGS in R (R-Core-Team, 2021). As the centers differed greatly in terms of species present, this analysis was performed separately for each center. Therefore, NGS data for each center were normalized with bestNormalized (Peterson, 2021) and then correlated with Pearson. The analysis was plotted in a heatmap using ggplot2 (Wickham, 2016). For each center, only the genera with the highest differentially abundance representing the six key players were selected based on Anova (p<0.001).

### Antibiotic resistance genes counts and virulence factors

MAGs and contigs were aligned against several databases in abricate (Seemann T, Abricate, Github https://github.com/tseemann/abricate) with options --mincov=70 and --minid=70, including HMO gene clusters sequences (Lawson et al., 2019), EcOH (Ingle et al., 2016), VFDB (Chen et al., 2005) and Resfinder (Zankari et al., 2012). To correlate antibiotic resistance gene patterns with specific genera, the taxonomy of the MAGs was assigned by GTDB-Tk (Chaumeil et al., 2020). The number of hits per infant and per time point is shown in a circle packing plot created with rawgraphs (Mauri et al., 2017). Data were analyzed on genus level whereas features were only depicted when the genus was represented by more than one MAG. Correlation of antibiotic resistance gene patterns with the six key species were visualized in a heatmap with R (R-Core-Team, 2021). All graphs were combined and assimilated in Inkscape v1.1 (URL: https://inkscape.org/en/ RRID:SCR_014479) to obtain a uniform appearance

## Supplemental Information

ASV-tables and metabolomic, as well as metabolomic data are shared via Github (https://github.com/CharlotteJNeumann/preterm_shared)

### 16.1) Supplemental information

**Suppl. Fig. 1:**
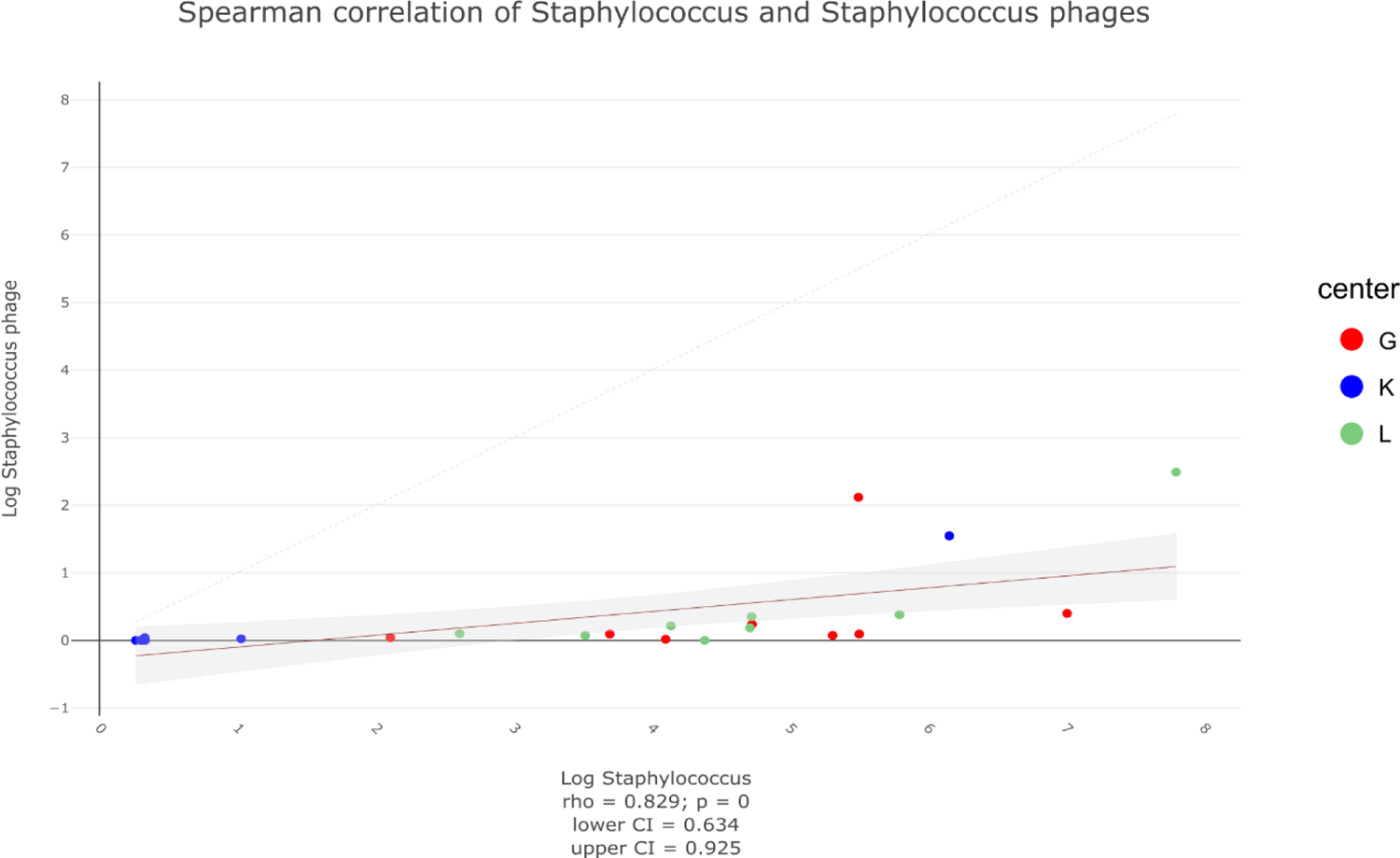
Spearman Correlation of Staphylococcus with Staphylococcus phages

**Suppl. Fig 2:**
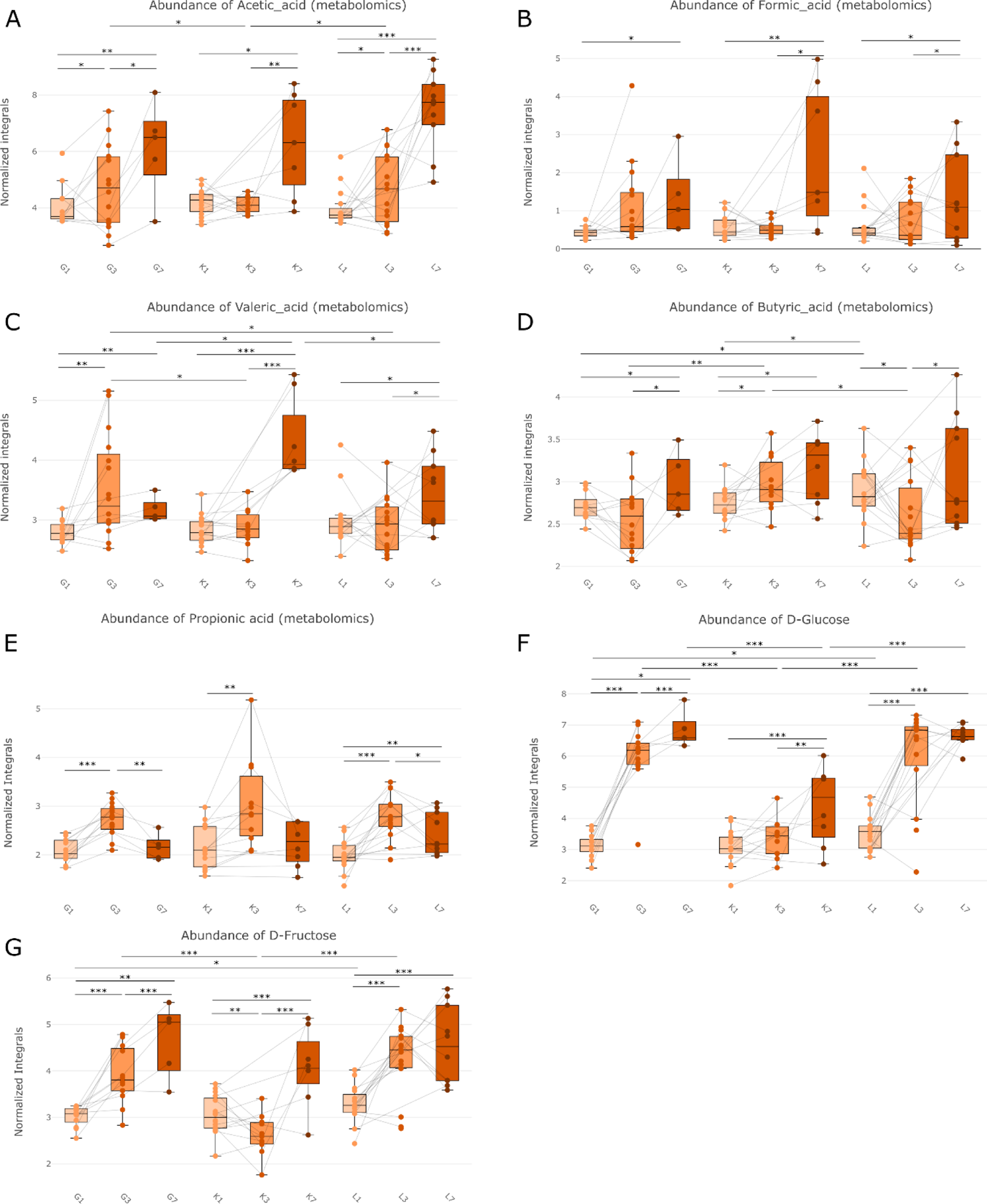
Normalized integrals of several metabolites over time (tp1, tp3, tp7) and centers (G, K, L); A) acetic acid, B) formic Acid, C) valeric acid, D) butyric acid, E) propionic acid, F) D-glucose, and G) D-fructose

**Suppl. Fig. 3:**
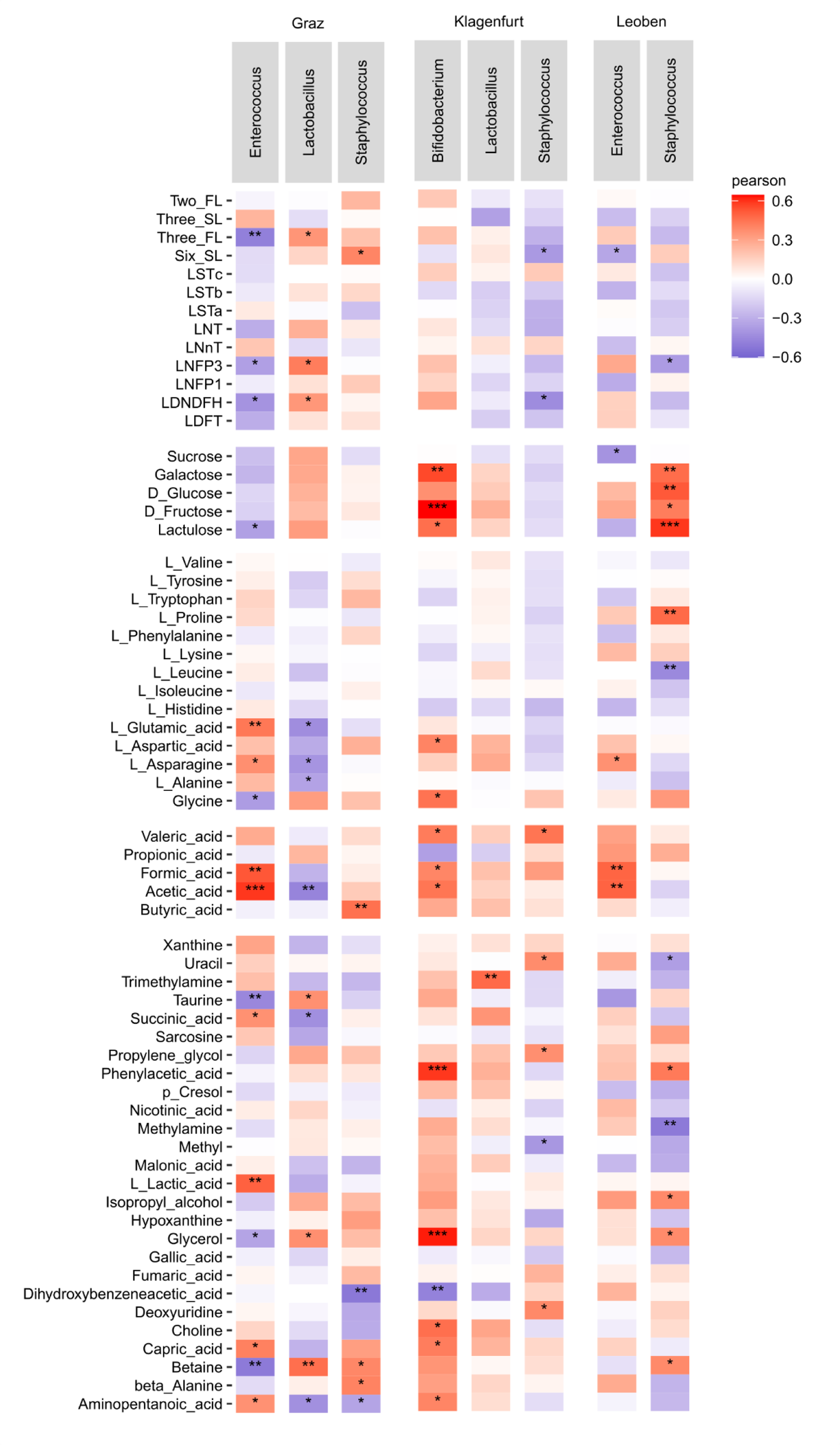
Complete heat map of correlation of metabolites and key-taxa of the three centers.

**Suppl. Table 1:** timepoint of stool sample collections

| Time Points | Minimum | Maximum | Mean |
| --- | --- | --- | --- |
| tp1 | 1 | 3 | 1.56 |
| tp2 | 3 | 6 | 3.75 |
| tp3 | 5 | 8 | 5.82 |
| tp4 | 7 | 15 | 8.04 |
| tp5 | 9 | 13 | 10.06 |
| tp6 | 11 | 15 | 12.15 |
| tp7 | 13 | 21 | 14.50 |

**Suppl. Table 2:** Bifidobacterium reference genomes

| sample | *Bifidobacterium longum* subsp. *longum* JCM1217<sup>T</sup> | *Bifidobacterium longum* subsp. *infantis* JCM1222<sup>T</sup> | *Bifidobacterium catenulatum* subsp. *Kashiwanohense* JCM15439<sup>T</sup> | *Bifidobacterium longum* subsp. *suillum* JCM19995<sup>T</sup> | *Bifidobacterium longum* subsp. *suis* JCM1269<sup>T</sup> | *Bifidobacterium breve* JCM1192<sup>T</sup> | *Bifidobacterium bifidum* JCM1255<sup>T</sup> |
| --- | --- | --- | --- | --- | --- | --- | --- |
| K08-7 | 46.59% | 86.80% | 3.99% | 49.90% | 48.67% | 17.49% | 3.70% |
| K11-7 | 50.14% | 97.98% | 4.31% | 53.85% | 52.17% | 18.70% | 3.81% |
| K13-7 | 52.26% | 95.56% | 4.22% | 55.66% | 54.27% | 19.55% | 4.20% |
| K14-7 | 53.31% | 00.07% | 4.28% | 56.91% | 55.43% | 19.80% | 4.15% |
| K15-7 | 51.31% | 97.60% | 4.30% | 54.76% | 53.22% | 19.36% | 4.09% |
| K16-7 | 3.07 % | 6.01% | 0.26% | 3.30% | 3.66% | 1.14% | 0.23% |
| K17-7 | 48.87% | 97.38% | 4.31% | 52.49% | 50.76% | 18.34% | 3.74% |
| K18-7 | 22.63% | 43.34% | 1.94% | 24.29% | 23.55% | 8.48% | 1.76% |

**Suppl. Table 3:**
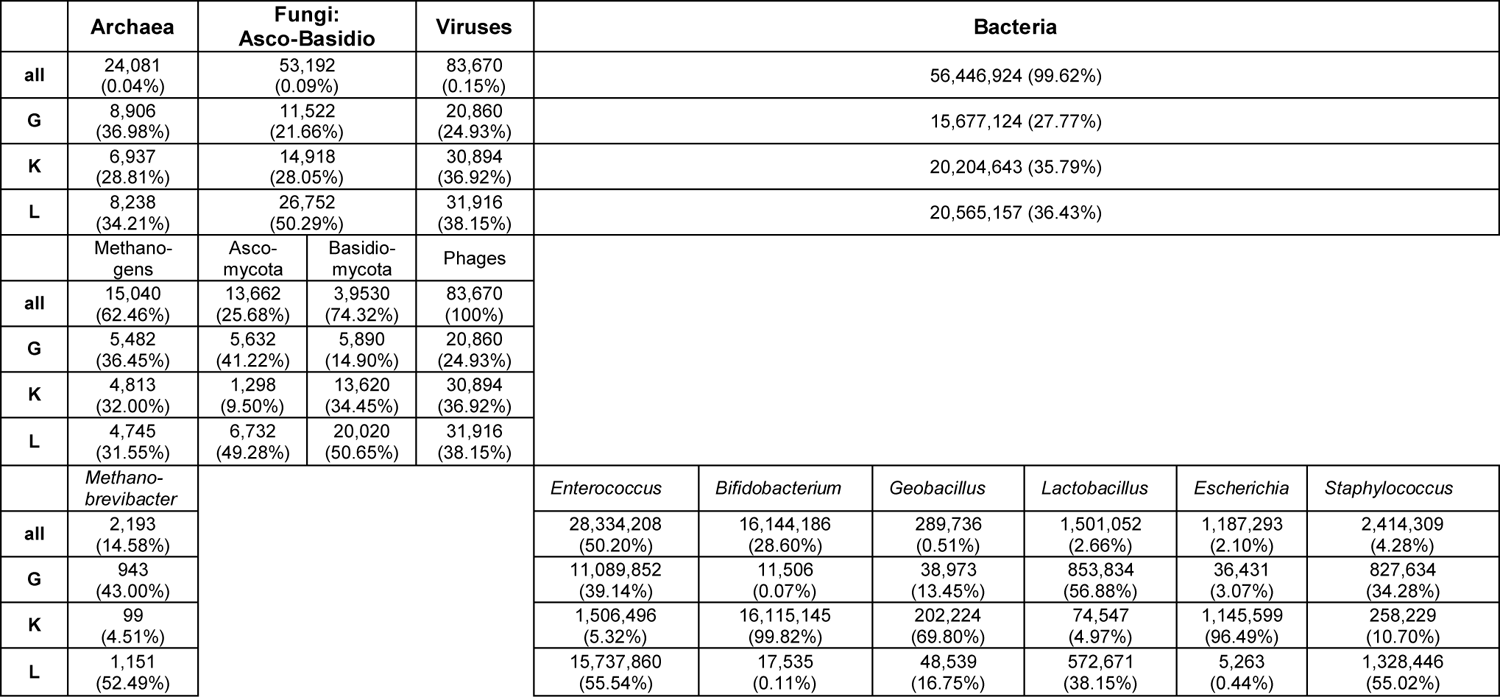
Distribution of overall reads (56,660,357) into Archaea, Ascomycetes-Basidiomycetes, Viruses and Bacteria as well as in their highest abundant subgroups. Distribution is given in overall reads and percentages and each for the three centers.

## Notes

### Competing Interest Statement

The authors have declared no competing interest.

### Author Declarations

Ethics committee of the Medical University of Graz gave ethical approval for this work (number 27-366 ex14/15) and written informed consent was obtained from the parents of the infants.

## References

1. Abdulkadir, B., Nelson, A., Skeath, T., Marrs, E.C.L., Perry, J.D., Cummings, S.P., Embleton, N.D., Berrington, J.E., and Stewart, C.J. (2016). Routine Use of Probiotics in Preterm Infants: Longitudinal Impact on the Microbiome and Metabolome. Neonatology 109, 239–247.

2. Alcon-Giner, C., Dalby, M.J., Caim, S., Ketskemety, J., Shaw, A., Sim, K., Lawson, M.A.E., Kiu, R., Leclaire, C., Chalklen, L., et al. (2020). Microbiota Supplementation with Bifidobacterium and Lactobacillus Modifies the Preterm Infant Gut Microbiota and Metabolome: An Observational Study. Cell Reports. Med. 1.

3. Alexander, V.N., Northrup, V., and Bizzarro, M.J. (2011). Antibiotic Exposure in the Newborn Intensive Care Unit and the Risk of Necrotizing Enterocolitis. J. Pediatr. 159, 392.

4. Alkan, H.F., Walter, K.E., Luengo, A., Madreiter-Sokolowski, C.T., Stryeck, S., Lau, A.N., Al-Zoughbi, W., Lewis, C.A., Thomas, C.J., Hoefler, G., et al. (2018). Cytosolic Aspartate Availability Determines Cell Survival When Glutamine Is Limiting. Cell Metab. 28, 706–720.e6.

5. Andrews S (2010). FastQC: a quality control tool for high throughput sequence data.

6. Austin, S., De Castro, C.A., Sprenger, N., Binia, A., Affolter, M., Garcia-Rodenas, C.L., Beauport, L., Tolsa, J.F., and Fumeaux, C.J.F. (2019). Human Milk Oligosaccharides in the Milk of Mothers Delivering Term versus Preterm Infants. Nutrients 11.

7. Bäckhed, F., Roswall, J., Peng, Y., Feng, Q., Jia, H., Kovatcheva-Datchary, P., Li, Y., Xia, Y., Xie, H., Zhong, H., et al. (2015). Dynamics and Stabilization of the Human Gut Microbiome during the First Year of Life. Cell Host Microbe 17, 690–703.

8. Bell, E.F. (2005). Preventing Necrotizing Enterocolitis: What Works and How Safe? Pediatrics 115, 173–174.

9. Birck, M.M., Nguyen, D.N., Cilieborg, M.S., Kamal, S.S., Nielsen, D.S., Damborg, P., Olsen, J.E., Lauridsen, C., Sangild, P.T., and Thymann, T. (2016). Enteral but not parenteral antibiotics enhance gut function and prevent necrotizing enterocolitis in formula-fed newborn preterm pigs. Am. J. Physiol. Gastrointest. Liver Physiol. 310, G323–G333.

10. Bizzarro, M.J. (2018). Avoiding Unnecessary Antibiotic Exposure in Premature Infants: Understanding When (Not) to Start and When to Stop. JAMA Netw. Open 1, e180165– e180165.

11. Blencowe, H., Cousens, S., Oestergaard, M.Z., Chou, D., Moller, A.B., Narwal, R., Adler, A., Vera Garcia, C., Rohde, S., Say, L., et al. (2012). National, regional, and worldwide estimates of preterm birth rates in the year 2010 with time trends since 1990 for selected countries: a systematic analysis and implications. Lancet (London, England) 379, 2162– 2172.

12. Bokulich, N.A., Kaehler, B.D., Rideout, J.R., Dillon, M., Bolyen, E., Knight, R., Huttley, G.A., and Gregory Caporaso, J. (2018). Optimizing taxonomic classification of marker-gene amplicon sequences with QIIME 2’s q2-feature-classifier plugin. Microbiome 6, 1–17.

13. Bolger, A.M., Lohse, M., and Usadel, B. (2014). Trimmomatic: a flexible trimmer for Illumina sequence data. Bioinformatics 30, 2114–2120.

14. Bolyen, E., Rideout, J.R., Dillon, M.R., Bokulich, N.A., Abnet, C.C., Al-Ghalith, G.A., Alexander, H., Alm, E.J., Arumugam, M., Asnicar, F., et al. (2019). Reproducible, interactive, scalable and extensible microbiome data science using QIIME 2. Nat. Biotechnol. 2019 378 37, 852–857.

15. Brown, C.T., Olm, M.R., Thomas, B.C., and Banfield, J.F. (2016). Measurement of bacterial replication rates in microbial communities. Nat. Biotechnol. 34, 1256–1263.

16. Brown, C.T., Olm, M.R., Thomas, B.C., Banfield, J.F., Biology, M., Science, P., Division, S., and Berkeley, L. (2017). Measurement of bacterial replication rates in microbial communities. Nat Biotechnol. 34, 1256–1263.

17. Buchfink, B., Reuter, K., and Drost, H.G. (2021). Sensitive protein alignments at tree-of-life scale using DIAMOND. Nat. Methods 2021 184 18, 366–368.

18. Bury, R.G., and Tudehope, D. (2001). Enteral antibiotics for preventing necrotizing enterocolitis in low birthweight or preterm infants. Cochrane Database Syst. Rev.

19. Callahan, B.J., McMurdie, P.J., Rosen, M.J., Han, A.W., Johnson, A.J.A., and Holmes, S.P. (2016). DADA2: High-resolution sample inference from Illumina amplicon data. Nat. Methods 2016 137 13, 581–583.

20. Chaumeil, P.A., Mussig, A.J., Hugenholtz, P., and Parks, D.H. (2020). GTDB-Tk: a toolkit to classify genomes with the Genome Taxonomy Database. Bioinformatics 36, 1925–1927.

21. Chen, L., Yang, J., Yu, J., Yao, Z., Sun, L., Shen, Y., and Jin, Q. (2005). VFDB: a reference database for bacterial virulence factors. Nucleic Acids Res. 33.

22. Corporation, B. (2012). Gentamicin(e) Product Monograph PRODUCT MONOGRAPH.

23. Costeloe, K., Bowler, U., Brocklehurst, P., Hardy, P., Heal, P., Juszczak, E., King, A., Panton, N., Stacey, F., Whiley, A., et al. (2016). A randomised controlled trial of the probiotic Bifidobacterium breve BBG-001 in preterm babies to prevent sepsis, necrotising enterocolitis and death: the Probiotics in Preterm infantS (PiPS) trial. Health Technol. Assess. 20, vii–83.

24. Dalby, M.J., and Hall, L.J. (2021). Populating preterm infants with probiotics. Cell Reports Med. 2, 100224.

25. Davis, N.M., Proctor, D.M., Holmes, S.P., Relman, D.A., and Callahan, B.J. (2018). Simple statistical identification and removal of contaminant sequences in marker-gene and metagenomics data. Microbiome 6, 226.

26. van Duin, D., and Paterson, D.L. (2016). Multidrug Resistant Bacteria in the Community: Trends and Lessons Learned. Infect. Dis. Clin. North Am. 30, 377.

27. Durand, L., Planchon, S., Guinebretiere, M.H., Carlin, F., and Remize, F. (2015). Genotypic and phenotypic characterization of foodborne Geobacillus stearothermophilus. Food Microbiol. 45, 103–110.

28. Esaiassen, E., Hjerde, E., Cavanagh, J.P., Pedersen, T., Andresen, J.H., Rettedal, S.I., Støen, R., Nakstad, B., Willassen, N.P., and Klingenberg, C. (2018). Effects of Probiotic Supplementation on the Gut Microbiota and Antibiotic Resistome Development in Preterm Infants. Front. Pediatr. 6, 347.

29. Fan, X., Zhang, L., Tang, J., Chen, C., Chen, J., Qu, Y., and Mu, D. (2017). The initial prophylactic antibiotic usage and subsequent necrotizing enterocolitis in high-risk premature infants: a systematic review and meta-analysis. Pediatr. Surg. Int. 2017 341 34, 35–45.

30. Fanaroff, A.A., Stoll, B.J., Wright, L.L., Carlo, W.A., Ehrenkranz, R.A., Stark, A.R., Bauer, C.R., Donovan, E.F., Korones, S.B., Laptook, A.R., et al. (2007). Trends in neonatal morbidity and mortality for very low birthweight infants. Am. J. Obstet. Gynecol. 196, 147.e1–147.e8.

31. Gustavsson, L., Lindquist, S., Elfvin, A., Hentz, E., and Studahl, M. (2020). Reduced antibiotic use in extremely preterm infants with an antimicrobial stewardship intervention. BMJ Paediatr. Open 4, e000872.

32. Högberg, N., Stenbäck, A., Carlsson, P.O., Wanders, A., and Lilja, H.E. (2013). Genes regulating tight junctions and cell adhesion are altered in early experimental necrotizing enterocolitis. J. Pediatr. Surg. 48, 2308–2312.

33. Ingle, D.J., Valcanis, M., Kuzevski, A., Tauschek, M., Inouye, M., Stinear, T., Levine, M.M., Robins-Browne, R.M., and Holt, K.E. (2016). In silico serotyping of E. coli from short read data identifies limited novel O-loci but extensive diversity of O:H serotype combinations within and between pathogenic lineages. Microb. Genomics 2.

34. Jin, Y.-T., Duan, Y., Deng, X.-K., and Lin, J. (2019). Prevention of necrotizing enterocolitis in premature infants – an updated review. World J. Clin. Pediatr. 8, 23.

35. Karthikeyan, G., and Bhat, B.V. (2017). The PiPS (Probiotics in Preterm Infants Study) Trial - Controlling the Confounding Factor of Cross-contamination Unveils Significant Benefits. Indian Pediatr. 54, 162.

36. Klopp, J., Ferretti, P., Meyer, C.U., Hilbert, K., Haiß, A., Marißen, J., Henneke, P., Hudalla, H., Pirr, S., Viemann, D., et al. (2022). Meconium Microbiome of Very Preterm Infants across Germany. MSphere.

37. Klymiuk, I., Bambach, I., Patra, V., Trajanoski, S., and Wolf, P. (2016). 16S based microbiome analysis from healthy subjects’ skin swabs stored for different storage periods reveal phylum to genus level changes. Front. Microbiol. 7, 2012.

38. Koskella, B., Hall, L.J., and Metcalf, C.J.E. (2017). The microbiome beyond the horizon of ecological and evolutionary theory. Nat. Ecol. Evol. 1, 1606–1615.

39. Kujawska, M., Collado, M.C., and Hall, L.J. (2021). Microbes, human milk, and prebiotics. Hum. Microbiome Early Life 197–237.

40. Kumar, M., Flint, S., Palmer, J., Chanapha, S., and Hall, C. (2021). Influence of Incubation Temperature and Total Dissolved Solids on Biofilm and Spore Formation by Dairy Isolates of Geobacillus stearothermophilus. Appl. Environ. Microbiol. 87, 1–10.

41. Kumpitsch, C., Fischmeister, F.P.S., Mahnert, A., Lackner, S., Wilding, M., Sturm, C., Springer, A., Madl, T., Holasek, S., Högenauer, C., et al. (2021). Reduced B12 uptake and increased gastrointestinal formate are associated with archaeome-mediated breath methane emission in humans. Microbiome 9, 1–18.

42. Kurath-Koller, S., Moissl-Eichinger, C., Gorkiewicz, G., Kraschl, R., Kanduth, C., Hopfer, B., Urlesberger, B., and Resch, B. (2017). Changes of intestinal microbiota composition and diversity in very low birth weight infants related to strategies of NEC prophylaxis: protocol for an observational multicentre pilot study. Pilot Feasibility Stud. 3, 52.

43. Kurath-Koller, S., Neumann, C., Moissl-Eichinger, C., Kraschl, R., Kanduth, C., Hopfer, B., Pausan, M.R., Urlesberger, B., and Resch, B. (2020). Hospital Regimens Including Probiotics Guide the Individual Development of the Gut Microbiome of Very Low Birth Weight Infants in the First Two Weeks of Life. Nutr. 2020, Vol. 12, Page 1256 *12*, 1256.

44. Langmead, B., and Salzberg, S.L. (2012). Fast gapped-read alignment with Bowtie 2. Nat. Methods 2012 94 9, 357–359.

45. Lawson, M.A.E., O’Neill, I.J., Kujawska, M., Gowrinadh Javvadi, S., Wijeyesekera, A., Flegg, Z., Chalklen, L., and Hall, L.J. (2019). Breast milk-derived human milk oligosaccharides promote Bifidobacterium interactions within a single ecosystem. ISME J. 2019 142 14, 635–648.

46. Li, D., Liu, C.M., Luo, R., Sadakane, K., and Lam, T.W. (2015). MEGAHIT: an ultra-fast single-node solution for large and complex metagenomics assembly via succinct de Bruijn graph. Bioinformatics 31, 1674–1676.

47. Li, H., Handsaker, B., Wysoker, A., Fennell, T., Ruan, J., Homer, N., Marth, G., Abecasis, G., and Durbin, R. (2009). The Sequence Alignment/Map format and SAMtools. Bioinformatics 25, 2078–2079.

48. Łubiech, K., and Twarużek, M. (2020). Lactobacillus Bacteria in Breast Milk. Nutrients 12, 1– 13.

49. Lyons, K.E., Ryan, C.A., Dempsey, E.M., Ross, R.P., and Stanton, C. (2020). Breast Milk, a Source of Beneficial Microbes and Associated Benefits for Infant Health. Nutr. 2020, Vol. 12, Page 1039 *12*, 1039.

50. Marcobal, A., and Sonnenburg, J.L. (2012). Human milk oligosaccharide consumption by intestinal microbiota. Clin. Microbiol. Infect. 18, 12.

51. Marcobal, A., Barboza, M., Froehlich, J.W., Block, D.E., German, J.B., Lebrilla, C.B., and Mills, D.A. (2010). Consumption of Human Milk Oligosaccharides by Gut-related Microbes. J. Agric. Food Chem. 58, 5334.

52. Martí, M., Spreckels, J.E., Ranasinghe, D., Sverremark-Ekströ, E., Jenmalm, M.C., Correspondence, T.A., Ranasinghe, P.D., Wejryd, E., Marchini, G., and Abrahamsson, T. (2021). Effects of Lactobacillus reuteri supplementation on the gut microbiota in extremely preterm infants in a randomized placebo-controlled trial. Cell Reports Med. 2, 100206.

53. Mauri, M., Elli, T., Caviglia, G., Uboldi, G., and Azzi, M. (2017). RAWGraphs: A visualisation platform to create open outputs. ACM Int. Conf. Proceeding Ser. Part F131371.

54. Meyer, F., Paarmann, D., D’Souza, M., Olson, R., Glass, E.M., Kubal, M., Paczian, T., Rodriguez, A., Stevens, R., Wilke, A., et al. (2008). The metagenomics RAST server - A public resource for the automatic phylogenetic and functional analysis of metagenomes. BMC Bioinformatics 9, 1–8.

55. Michael Cotten, C., Taylor, S., Stoll, B., Goldberg, R.N., Hansen, N.I., Sanchez, P.J., Ambalavanan, N., and Benjamin, D.K. (2009). Prolonged Duration of Initial Empirical Antibiotic Treatment Is Associated With Increased Rates of Necrotizing Enterocolitis and Death for Extremely Low Birth Weight Infants. Pediatrics 123, 58.

56. Moissl-Eichinger, C., Probst, A.J., Birarda, G., Auerbach, A., Koskinen, K., Wolf, P., and Holman, H.Y.N. (2017). Human age and skin physiology shape diversity and abundance of Archaea on skin. Sci. Rep. 7.

57. Nazina, T.N., Tourova, T.P., Poltaraus, A.B., Novikova, E. V., Grigoryan, A.A., Ivanova, A.E., Lysenko, A.M., Petrunyaka, V. V., Osipov, G.A., Belyaev, S.S., et al. (2001). Taxonomic study of aerobic thermophilic bacilli: Descriptions of Geobacillus subterraneus gen. nov., sp. nov. and Geobacillus uzenensis sp. nov. from petroleum reservoirs and transfer of Bacillus stearothermophilus, Bacillus thermocatenulatus, Bacillus thermoleovorans. Int. J. Syst. Evol. Microbiol. 51, 433–446.

58. Neu, J., and Walker, W.A. (2011). Necrotizing Enterocolitis. N. Engl. J. Med. 364, 255.

59. Oki, K., Akiyama, T., Matsuda, K., Gawad, A., Makino, H., Ishikawa, E., Oishi, K., Kushiro, A., and Fujimoto, J. (2018). Long-term colonization exceeding six years from early infancy of Bifidobacterium longum subsp. longum in human gut. BMC Microbiol. 18, 1–13.

60. Oksanen, A.J., Blanchet, F.G., Kindt, R., Legendre, P., Minchin, P.R., Hara, R.B.O., Simpson, G.L., Solymos, P., Stevens, M.H.H., and Wagner, H. (2015). The vegan package. Community ecology package. Http://CRAN.R-Project.Org/Package=vegan.

61. Olm, M.R., Brown, C.T., Brooks, B., and Banfield, J.F. (2017). dRep: a tool for fast and accurate genomic comparisons that enables improved genome recovery from metagenomes through de-replication. ISME J. 2017 1112 11, 2864–2868.

62. Ondov, B.D., Bergman, N.H., and Phillippy, A.M. (2011). Interactive metagenomic visualization in a Web browser. BMC Bioinformatics 12, 1–10.

63. Overbeek, R., Begley, T., Butler, R.M., Choudhuri, J. V., Chuang, H.Y., Cohoon, M., de Crécy-Lagard, V., Diaz, N., Disz, T., Edwards, R., et al. (2005). The subsystems approach to genome annotation and its use in the project to annotate 1000 genomes. Nucleic Acids Res. 33, 5691–5702.

64. Parks, D.H., Imelfort, M., Skennerton, C.T., Hugenholtz, P., and Tyson, G.W. (2015). CheckM: assessing the quality of microbial genomes recovered from isolates, single cells, and metagenomes. Genome Res. 25, 1043–1055.

65. Pausan, M.R., Csorba, C., Singer, G., Till, H., Schöpf, V., Santigli, E., Klug, B., Högenauer, C., Blohs, M., and Moissl-Eichinger, C. (2019). Exploring the Archaeome: Detection of Archaeal Signatures in the Human Body. Front. Microbiol. 10, 2796.

66. Peterson, R.A. (2021). Finding Optimal Normalizing Transformations via bestNormalize. R J. 13, 310--329.

67. Picard, C., Fioramonti, J., Francois, A., Robinson, T., Neant, F., and Matuchansky, C. (2005). Review article: bifidobacteria as probiotic agents – physiological effects and clinical benefits. Aliment. Pharmacol. Ther. 22, 495–512.

68. Probst, A.J., Auerbach, A.K., and Moissl-Eichinger, C. (2013). Archaea on Human Skin. PLoS One 8, e65388.

69. Quast, C., Pruesse, E., Yilmaz, P., Gerken, J., Schweer, T., Yarza, P., Peplies, J., and Glöckner, F.O. (2013). The SILVA ribosomal RNA gene database project: improved data processing and web-based tools. Nucleic Acids Res. 41.

70. Quinlan, A.R., and Hall, I.M. (2010). BEDTools: a flexible suite of utilities for comparing genomic features. Bioinformatics 26, 841–842.

71. R-Core-Team, R. (2021). R: A Language and Environment for Statistical Computing.

72. Ramsey, M., Hartke, A., and Huycke, M. (2014). The Physiology and Metabolism of Enterococci. Enterococci From Commensals to Lead. Causes Drug Resist. Infect.

73. Reeder, J., Huang, M., Kaminker, J.S., and Paulson, J.N. (2021). MicrobiomeExplorer: an R package for the analysis and visualization of microbial communities. Bioinformatics 37, 1317.

74. Ruiz, L., García-Carral, C., and Rodriguez, J.M. (2019). Unfolding the human milk microbiome landscape in the omicsera. Front. Microbiol. 10, 1378.

75. Schmolzer, G., Urlesberger, B., Haim, M., Kutschera, J., Pichler, G., Ritschl, E., Resch, B., Reiterer, F., and Müller, W. (2006). Multi-modal approach to prophylaxis of necrotizing enterocolitis: Clinical report and review of literature. Pediatr. Surg. Int. 22, 573–580.

76. Soto, A., Martín, V., Jiménez, E., Mader, I., Rodríguez, J.M., and Fernández, L. (2014). Lactobacilli and Bifidobacteria in Human Breast Milk: Influence of Antibiotherapy and Other Host and Clinical Factors. J. Pediatr. Gastroenterol. Nutr. 59, 78.

77. Turenne, C.Y., Sanche, S.E., Hoban, D.J., Karlowsky, J.A., and Kabani, A.M. (1999). Rapid Identification of Fungi by Using the ITS2 Genetic Region and an Automated Fluorescent Capillary Electrophoresis System. J. Clin. Microbiol. 37, 1846.

78. Underwood, M.A., and Sohn, K. (2017). The Microbiota of the Extremely Preterm Infant. Clin. Perinatol. 44, 407.

79. Wang, X., Li, J., Li, N., Guan, K., Yin, D., Zhang, H., Ding, G., and Hu, Y. (2021). Evolution of Intestinal Gases and Fecal Short-Chain Fatty Acids Produced in vitro by Preterm Infant Gut Microbiota During the First 4 Weeks of Life. Front. Pediatr. 0, 1011.

80. Wellmann, F. (2018). Epidemiologie der nekrotisierenden Enterokolitis im südlichen Österreich eine retrospektive Studie.

81. White, T.J., Bruns, T., Lee, S., and Taylor, J. (1990). Amplification and direct sequencing of fungal ribosomal RNA genes for phylogenetics. PCR Protoc. 315–322.

82. Wickham, H. (2016). ggplot2: Elegant Graphics for Data Analysis.

83. World Health Organization (2015). Global action plan on antimicrobial resistance.

84. Wu, Y.W., Tang, Y.H., Tringe, S.G., Simmons, B.A., and Singer, S.W. (2014). MaxBin: An automated binning method to recover individual genomes from metagenomes using an expectation-maximization algorithm. Microbiome 2, 1–18.

85. Zakrzewski, M., Proietti, C., Ellis, J.J., Hasan, S., Brion, M.J., Berger, B., and Krause, L. (2017). Calypso: a user-friendly web-server for mining and visualizing microbiome– environment interactions. Bioinformatics 33, 782.

86. Zankari, E., Hasman, H., Cosentino, S., Vestergaard, M., Rasmussen, S., Lund, O., Aarestrup, F.M., and Larsen, M.V. (2012). Identification of acquired antimicrobial resistance genes. J. Antimicrob. Chemother. 67, 2640–2644.

87. Zheng, J., Wittouck, S., Salvetti, E., Franz, C.M.A.P., Harris, H.M.B., Mattarelli, P., O’Toole, P.W., Pot, B., Vandamme, P., Walter, J., et al. (2020). A taxonomic note on the genus Lactobacillus: Description of 23 novel genera, emended description of the genus Lactobacillus Beijerinck 1901, and union of Lactobacillaceae and Leuconostocaceae. Int. J. Syst. Evol. Microbiol. 70, 2782–2858.

